# Determinants of antibody responses to two doses of ChAdOx1 nCoV-19 or BNT162b2 and a subsequent booster dose of BNT162b2 or mRNA-1273: population-based cohort study (COVIDENCE UK)

**DOI:** 10.1101/2022.02.14.22270930

**Authors:** David A Jolliffe, Sian E Faustini, Hayley Holt, Natalia Perdek, Sheena Maltby, Mohammad Talaei, Matthew Greenig, Giulia Vivaldi, Florence Tydeman, Jane Symons, Gwyneth A Davies, Ronan A Lyons, Christopher J Griffiths, Frank Kee, Aziz Sheikh, Seif O Shaheen, Alex G Richter, Adrian R Martineau

## Abstract

**Background:** Antibody responses to SARS-CoV-2 vaccination vary for reasons that remain poorly understood.

**Methods:** We tested for presence of combined IgG, IgA and IgM (IgGAM) anti-spike antibodies before and after administration of two doses of ChAdOx1 nCoV-19 (ChAdOx1, Oxford-AstraZeneca) or BNT162b2 (Pfizer-BioNTech) in UK adults participating in a population-based longitudinal study who received their first dose of vaccine from December 15, 2020 to July 10, 2021. Information on sixty-six potential sociodemographic, behavioural, clinical, pharmacological and nutritional determinants of serological response to vaccination was captured using serial online questionnaires. We used logistic regression to estimate multivariable-adjusted odds ratios (aORs) for associations between independent variables and risk of seronegativity following two vaccine doses. Participants who were seronegative after receiving two vaccine doses were offered an additional antibody test following subsequent administration of a ‘booster’ dose of BNT162b2 or mRNA-1273 (Moderna) from September 23 to December 12, 2021.

**Findings:** Serology results following two vaccine doses were available for 9,101 participants, of whom 5,770 (63.4%) received ChAdOx1 and 3,331 (36.6%) received BNT162b2. Anti-spike IgGAM was undetectable in 378 (4.2%) participants at a median of 8.6 weeks (IQR 6.4-10.7 weeks) after their second dose of vaccine. Seronegativity following two doses of SARS-CoV-2 vaccination was associated with administration of ChAdOx1 *vs* BNT162b2 (aOR 7.03, 95% CI 4.39-11.24), shorter interval between first and second vaccine doses (aOR 2.37, 1.06-5.26, for <6 weeks *vs* >10 weeks; aOR 1.59, 1.18-2.13, for 6-10 weeks *vs* >10 weeks), poorer self-assessed general health (aOR 3.33, 1.49-7.46, for poor *vs* excellent), immunodeficiencies (aOR 6.75, 2.63-17.35) and prescription of systemic immunosuppressants (aOR 3.76, 2.44-5.78). By contrast, pre-vaccination SARS-CoV-2 seropositivity (aOR 0.16, 0.04-0.70, for symptomatic seropositives *vs* seronegatives) and supplemental vitamin D intake (aOR 0.73, 0.53-0.99) were associated with reduced risk of post-vaccination seronegativity. 247/378 (65.3%) of participants who were seronegative after two doses of ChAdOx1 *vs* BNT162b2 provided a third sample at a median of 7.8 weeks (IQR 5.8-10.4) after receiving a booster dose of BNT162b2 or mRNA-1273: eight (3.2%) of them remained seronegative after three vaccine doses, all of whom either had a primary immunodeficiency or were taking systemic immunosuppressant drugs.

**Interpretation:** We identify multiple determinants of antibody responses to two doses of ChAdOx1 or BNT162b2, many of which are potentially modifiable. Booster doses of BNT162b2 or mRNA-1273 were highly effective in achieving seroconversion in those who failed to mount antibody responses following two doses of ChAdOx1 or BNT162b2.

**Study registration:** https://clinicaltrials.gov/ct2/show/NCT04330599

**Funding:** Barts Charity, Fischer Family Trust, The Exilarch’s Foundation, DSM Nutritional Products, Health Data Research UK

**Research in context:** *Evidence before this study:* We searched PubMed, medRxiv, and Google Scholar for papers published from January 1, 2020, to February 1, 2022, using the search terms (antibody OR humoral OR serologic* OR immunogenic*) AND (SARS-CoV-2 vaccine OR ChAdOx1 or BNT162b2 coronavirus), with no language restrictions. Population-based studies investigating multiple potential determinants of vaccine immunogenicity in people with known pre-vaccination SARS-CoV-2 serostatus are lacking.

*Added value of this study:* This large population-based study, conducted in a population with known pre-vaccination SARS-CoV-2 serostatus, examines a comprehensive range of potential sociodemographic, behavioural, clinical, pharmacological and nutritional determinants of antibody responses to administration of two major SARS-CoV-2 vaccines (i.e., ChAdOx1 or BNT162b2), many of which have not previously been investigated. It is also the first population-based study to characterise antibody responses to booster doses of SARS-CoV-2 vaccines in adults who were seronegative after their primary course of vaccination.

*Implications of all the available evidence:* Increased risk of seronegativity following two doses of SARS-CoV-2 vaccines was associated with administration of ChAdOx1 *vs* BNT162b2, shorter interval between first and second vaccine doses, poorer self-assessed general health, immunocompromise and SARS-CoV-2 seronegativity pre-vaccination. Regular intake of vitamin D supplements was associated with reduced risk of post-vaccination seronegativity. Randomised controlled trials are now needed to test for causality. Booster doses of BNT162b2 or mRNA-1273 were highly effective in achieving seroconversion in the majority of people who failed to mount antibody responses following a primary course of vaccination, the few exceptions being a subset of those with primary immunodeficiency or systemic immunosuppressant drugs.

## Introduction

Vaccination against SARS-CoV-2 represents a key tool for COVID-19 control. However, existing vaccines offer imperfect protection against disease, reflecting heterogeneity in immunogenicity.^1,2^ Improved understanding of factors influencing immune responses to SARS-CoV-2 vaccines could lead to discovery of effect-modifiers that could be harnessed to augment vaccine immunogenicity, and identify groups of poor responders who might benefit from more intensive vaccine dosing regimens or implementation of other protective measures.^3^

Existing studies investigating determinants of vaccine immunogenicity have reported that lower antibody responses following SARS-CoV-2 vaccination associate with administration of viral vector *vs* messenger RNA (mRNA) vaccines, older age, poorer general health, immunosuppression and shorter inter-dose intervals.^4-11^ Higher post-vaccination antibody titres are seen in those with evidence of SARS-CoV-2 infection before vaccination.^9,12,13^ However, these studies are limited in several respects: many are conducted in specific populations such as health care workers^5,9,12,13^ or in individuals with a particular demographic or clinical characteristic that may influence vaccine immunogenicity,^6,10,11^ which may constrain generalisability of their results. Where population-based studies have been done,^4,7,8^ these did not compare the effect of two doses of viral vector *vs* mRNA vaccines on host response; neither did they systematically ascertain participants’ pre-vaccination SARS-CoV-2 serostatus, which is an important potential confounder of associations reported. Moreover, these studies did not investigate effects of modifiable factors that have been posited to influence responses to vaccination, such as time of day of inoculation,^14^ nutrition,^15^ sleep,^16^ alcohol use,^17^ tobacco smoking^18^ and peri-vaccination use of anti-pyretic analgesics.^19^ There is also a lack of population-based studies investigating responses to booster doses of SARS-CoV-2 vaccines in individuals who failed to mount an antibody response after their primary course of vaccination.

We sought to address these limitations by investigating a comprehensive range of potential sociodemographic, behavioural, clinical, pharmacological and nutritional determinants of antibody responses in a population-based cohort of UK adults (COVIDENCE UK)^20^ following administration of two doses of either BNT162b2 (Pfizer-BioNTech) or ChAdOx1 nCoV-19 (Oxford-AstraZeneca; hereafter ChAdOx1) SARS-CoV-2 vaccines. Post-booster tests were offered to participants who were seronegative after their primary course of vaccination. We utilised an assay with proven sensitivity for detection of combined IgG, IgA and IgM antibodies to the SARS-CoV-2 spike antigen^21^ that has been validated as a correlate of protection against breakthrough COVID-19 in two populations.^22,23^

## Methods

### Study design and participants

COVIDENCE UK is a prospective, longitudinal, population-based observational study of COVID-19 in the UK population (www.qmul.ac.uk/covidence).^20^ Inclusion criteria were age ≥16 years and UK residence at enrolment, with no exclusion criteria. Participants were invited *via* a national media campaign to complete an online baseline questionnaire to capture information on potential symptoms of COVID-19 experienced since February 1, 2020; results of any COVID-19 tests; and details of a wide range of potential risk factors for COVID-19 and determinants of vaccine response (Table S1, Supplementary Appendix). Online monthly follow-up questionnaires captured incident test-confirmed COVID-19 and vaccination details (Table S2, Supplementary Appendix), and a pre-vaccination serology study was conducted to evaluate risk factors for SARS-CoV-2 infection.^24^ The study was launched on May 1, 2020, and closed to enrolment on October 6, 2021. Participants in the cohort were invited *via* email to participate in post-vaccination antibody studies and to give additional consent for these. The primary analysis presented here includes data from all participants for whom a serology result was available following administration of two doses of ChAdOx1 or BNT162b2; an additional antibody test following administration of a third booster dose of vaccine was offered to all participants who were seronegative after two vaccine doses, and a randomly selected subset of positive controls who were seropositive after two vaccine doses.

COVIDENCE UK was sponsored by Queen Mary University of London and approved by Leicester South Research Ethics Committee (ref 20/EM/0117). It is registered with ClinicalTrials.gov (NCT04330599).

### Procedures

Participants were sent kits containing instructions, lancets, and blood spot collection cards, to be posted back to the study team. Once returned, the samples were logged by the study team and sent in batches to the Clinical Immunology Service at the Institute of Immunology and Immunotherapy of the University of Birmingham (Birmingham, UK) for semi-quanttative determination of combined IgG, IgM and IgA anti-spike antibody titres using a commercially available ELISA that measures combined IgG, IgA, and IgM (IgGAM) responses to the SARS-CoV-2 trimeric spike glycoprotein (product code MK654; The Binding Site [TBS], Birmingham, UK). This assay has been CE-marked with 98.3% (95% Confidence Interval [CI] 96.4–99.4) specificity and 98.6% (92.6–100.0) sensitivity following reverse transcription polymerase chain reaction (RT-PCR)-confirmed mild-to-moderate COVID-19 that did not result in hospitalisation.^25^ A cut-off ratio relative to the TBS cut-off calibrators was determined by plotting 624 pre-2019 negatives in a frequency histogram. A cut-off coefficient was then established for IgGAM (1.31), with ratio values classed as positive (≥1) or negative (<1). Dried blood spot eluates were pre-diluted 1:40 with 0.05% PBS-Tween using a Dynex Revelation automated absorbance microplate reader (Dynex Technologies; Chantilly, VA, USA). Plates were developed after 10 min using 3,3′,5,5′-tetramethylbenzidine core, and orthophosphoric acid used as a stop solution (both TBS). Optical densities at 450 nm were measured using the Dynex Revelation.

### Outcomes

Study outcomes were presence *vs* absence of antibodies against SARS-CoV-2 (analysed as a binary outcome) and antibody titres (continuous outcome restricted to the individuals who were seropositive after receiving two vaccine doses).

### Independent variables

Sixty-six putative determinants of post-vaccination serological responses were selected *a priori*, covering vaccination type and timing, sociodemographic, occupational, and lifestyle factors; longstanding medical conditions and prescribed medication use; Bacille Calmette Guérin (BCG) vaccine status, and diet and supplemental micronutrient intake (Tables S1-2, Supplementary Appendix). These factors, which were obtained from the baseline or monthly questionnaires, were included as independent variables in our models. To produce participant-level covariates for each class of medications investigated, questionnaire responses were mapped to drug classes listed in the British National Formulary (BNF) or the DrugBank and Electronic Medicines Compendium databases if not explicitly listed in the BNF, as previously described.^20^ Index of Multiple Deprivation (IMD) 2019 scores were assigned according to participants’ postcodes, and categorised into quartiles.

### Statistical analysis

Full details of the sample size calculation and statistical analysis are provided in Supplementary Material. Briefly, logistic regression models were used to estimate adjusted ORs, 95% (CIs) and associated pairwise P-values for potential determinants of post-vaccination seronegativity in all participants. Linear regression models were used to estimate beta-coefficients, 95% CIs and associated pairwise P-values for potential determinants of log-transformed antibody titres in the subset of seropositive participants. For ease of interpretation, log-transformed estimates of antibody titres were exponentiated and a percentage increase or decrease was calculated for every one-unit increase in the potential determinant. We first estimated ORs and beta-coefficients in minimally adjusted models, and included factors independently associated with each outcome at the 10% significance level in fully adjusted models.

In a sensitivity analysis, we excluded participants from the analysis of antibody titres after two vaccine doses who reported a positive RT-PCR or lateral flow test for SARS-CoV-2 between the date of their second dose of vaccine and the date on which they provided their dried blood spot sample. We also stratified the analysis of antibody titres following two vaccine doses according to the type of vaccine received to explore whether determinants of antibody responses to two vaccine doses were consistent for ChAdOx1 *vs* BNT162b2. Given reports that peri-vaccination use of antipyretic analgesics may attenuate vaccine immunogenicity,^26^ we also conducted an exploratory analysis to determine the influence of taking paracetamol or non-steroidal anti-inflammatory drugs (NSAIDs) to treat post-vaccination symptoms on post-vaccination antibody titres.

## Results

A total of 15,527 participants were invited to participate in the study of antibody responses following two vaccine doses, of whom 13,005 consented to take part. Serology results were available for 9,244 participants, of whom 9,101 received two doses of ChAdOx1 (n=5,770) or BNT162b2 (n=3,331) for their primary vaccination course and were included in this analysis (Figure S1, Supplementary Appendix). Their characteristics are presented in Table 1: median age was 64.2 years at enrolment (IQR 57.1, 69.9), 71.1% were female, and 96.5% were of White ethnic origin. Post-vaccination dried blood spot samples were provided from January 12 to September 18, 2021 by participants who received their first dose of vaccine from December 15, 2020 to July 10, 2021 and their second dose from January 5, 2021 to September 4, 2021. Median time from the date of participants’ second vaccine dose to the date on which post-vaccination dried blood spot samples were provided was 8.6 weeks (IQR 6.4-10.7 weeks) and 374 of 9,101 participants (4.1%) were seronegative following two doses of vaccine. All of these participants, along with a randomly selected subset of 100 positive controls (who were seropositive after two doses of vaccine) were offered a third antibody test following administration of a single booster vaccine dose between September 23 to December 12, 2021. A total of 304 of these participants provided post-booster dried blood spot samples. Their characteristics are presented in Table S3: 261 received a BNT162b2 booster (221 after two doses of ChAdOx1 and 40 after two doses of BNT162b2) and 43 received a mRNA-1273 (Moderna) booster (all after two doses of ChAdOx1).

**Table 1.**
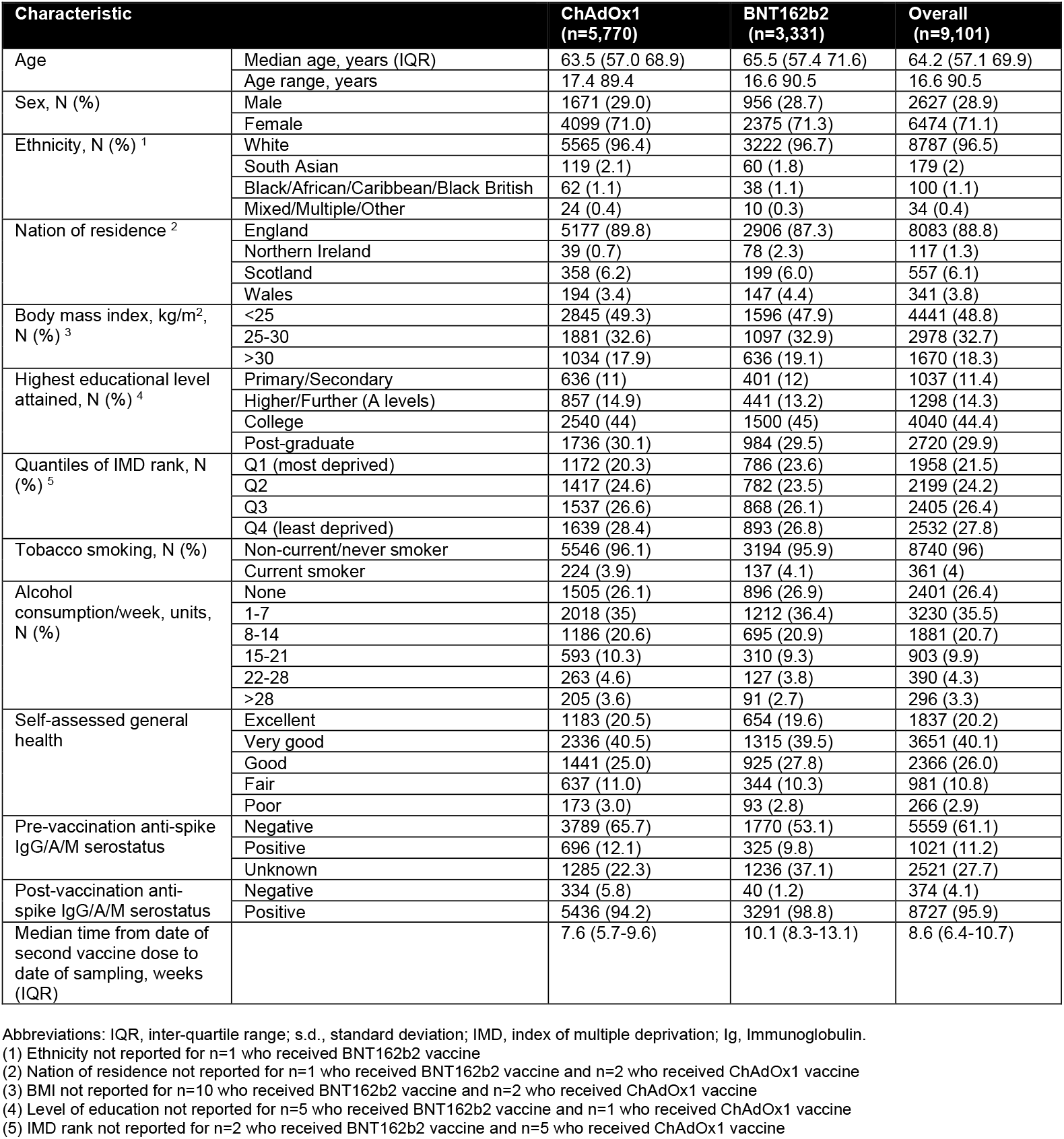
Participant characteristics, by vaccine type and overall (n=9,101)

### Determinants of seronegativity following two vaccine doses

Table 2 presents results of minimally adjusted and multivariable analyses to identify factors that were independently associated with the absence of detectable anti-spike IgG/A/M following two doses of ChAdOx1 or BNT162b2. After adjusting for age and sex, 32 independent variables associated with post-vaccination serostatus with P<0.10 and were fitted in a fully adjusted model. Ten factors in the fully adjusted model remained independently associated with post-vaccination serostatus (P<0.05). Eight of these were associated with higher risk of post-vaccination seronegativity: receiving ChAdOx1 *vs* BNT162b2 vaccine (aOR 7.03, 95% CI 4.39-11.24); shorter interval between first and second vaccine doses (aOR 1.59, 1.18-2.13, for 6-10 weeks *vs* >10 weeks, and aOR 2.37, 1.06-5.26, for <6 weeks *vs* >10 weeks); older age (aOR per 10-year increase in age 1.68, 1.42-2.00), poorer self-assessed general health (aOR per level of declining health, 1.26, 1.09-1.45); immunodeficiency disorder (aOR 6.75, 2.63-17.35), and use of systemic immunosuppressants (aOR 3.76, 2.44-5.78), sodium-glucose co-transporter-2 inhibitors (aOR 3.75, 1.12-12.52) and anti-platelet agents (aOR 2.87, 1.08-7.64). Two factors were associated with lower risk of post-vaccination seronegativity: pre-vaccination seropositivity for SARS-CoV-2 (aOR 0.54, 0.32-0.90, for individuals who were seropositive without COVID-19 symptoms pre-vaccination *vs* those who were seronegative pre-vaccination, and aOR 0.16, 0.04-0.70, for individuals who were seropositive with COVID-19 symptoms pre-vaccination *vs* those who were seronegative pre-vaccination) and regular consumption of vitamin D supplements (aOR 0.68, 0.52-0.89). No associations were seen for other independent variables investigated, including time of day of inoculation, nutrition, sleep, alcohol use, tobacco smoking or socioeconomic deprivation.

**Table 2.**
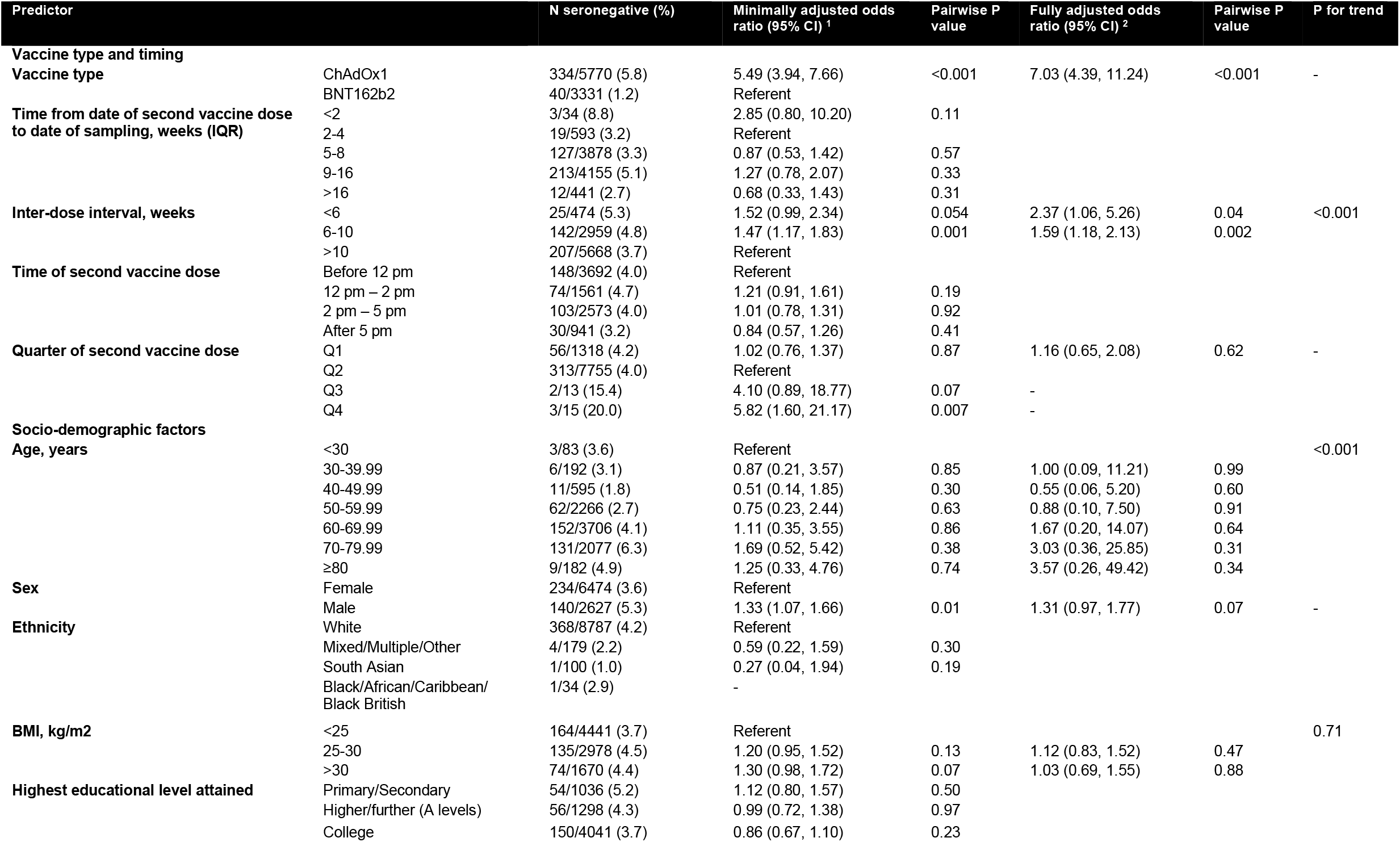
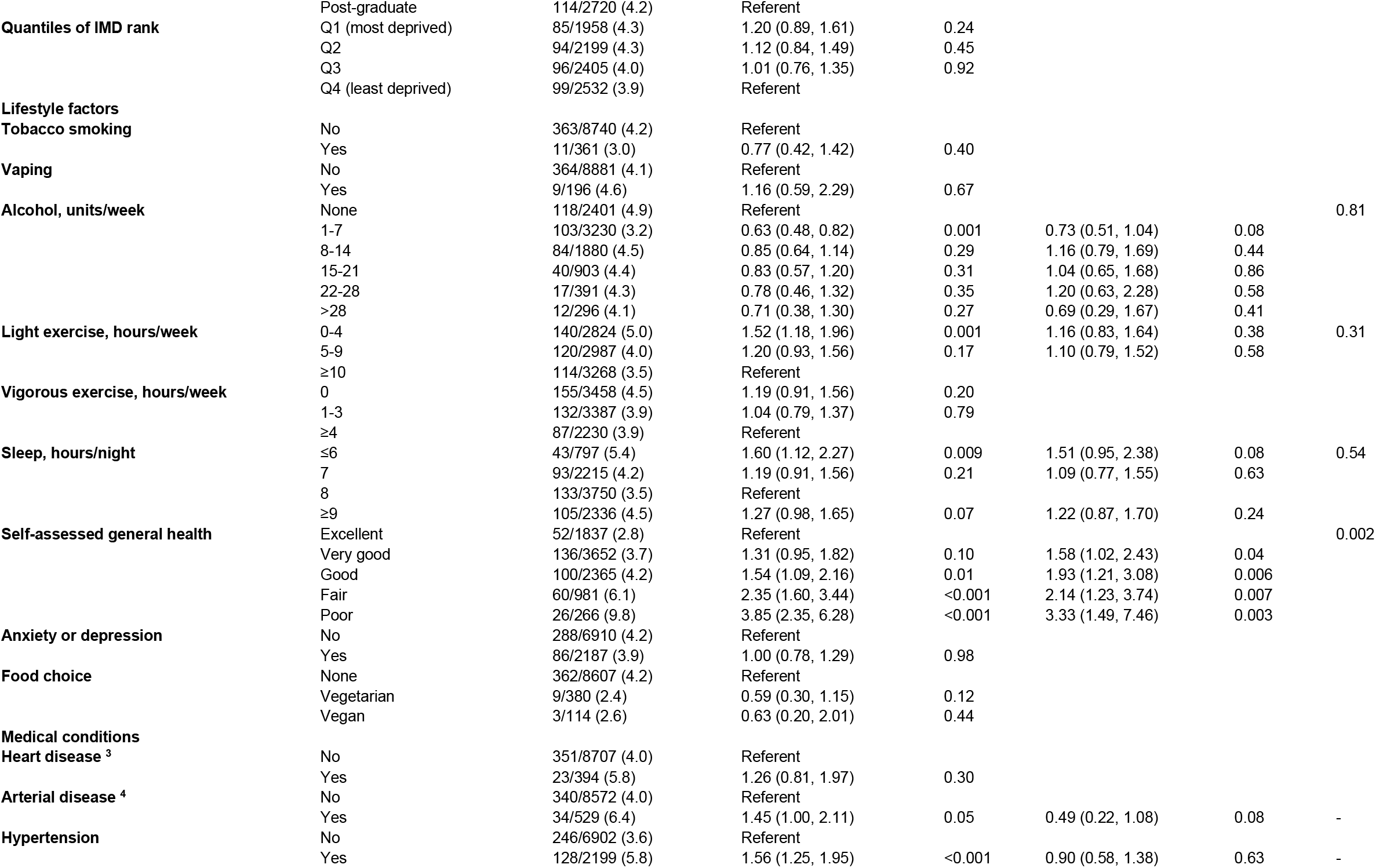
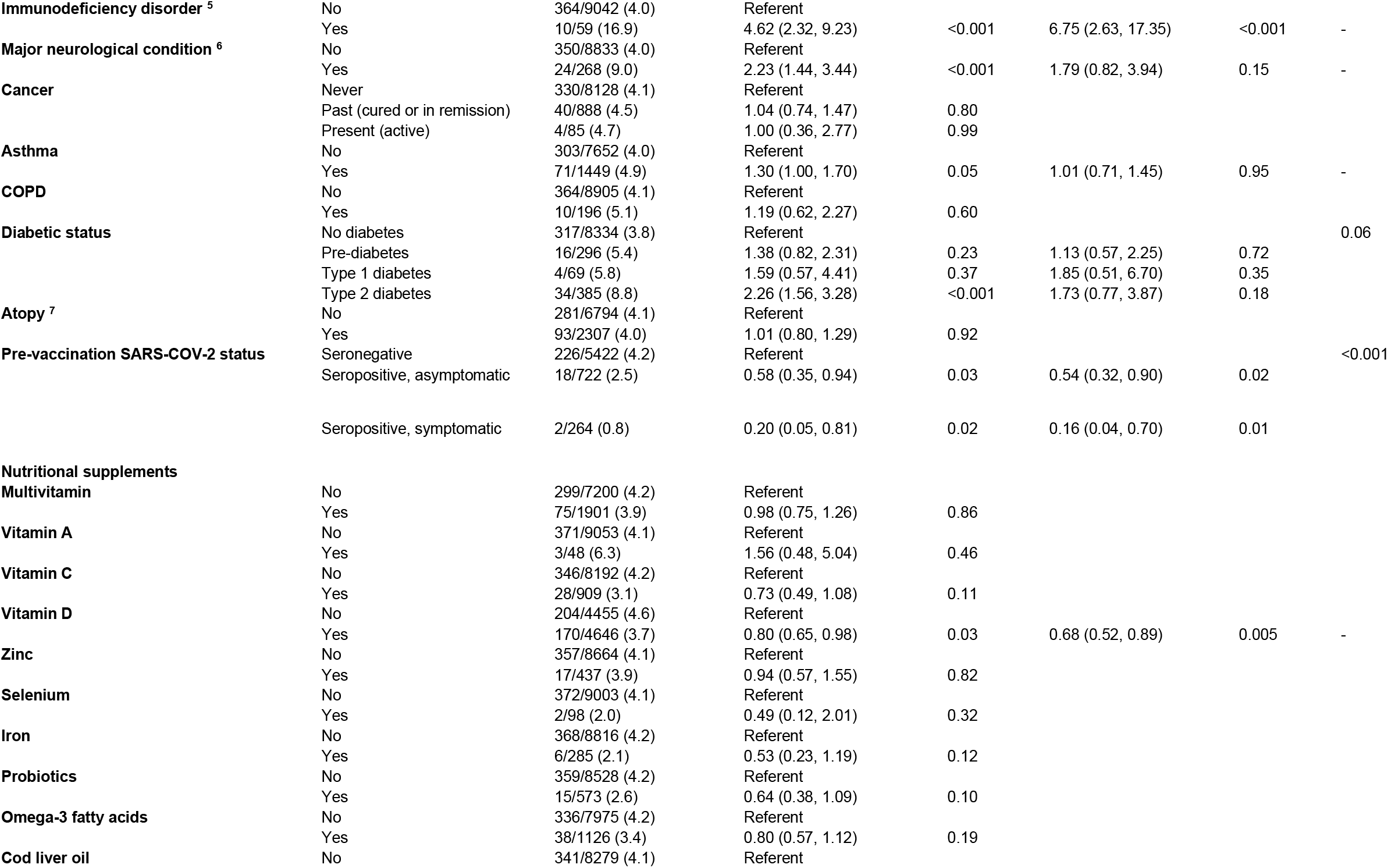
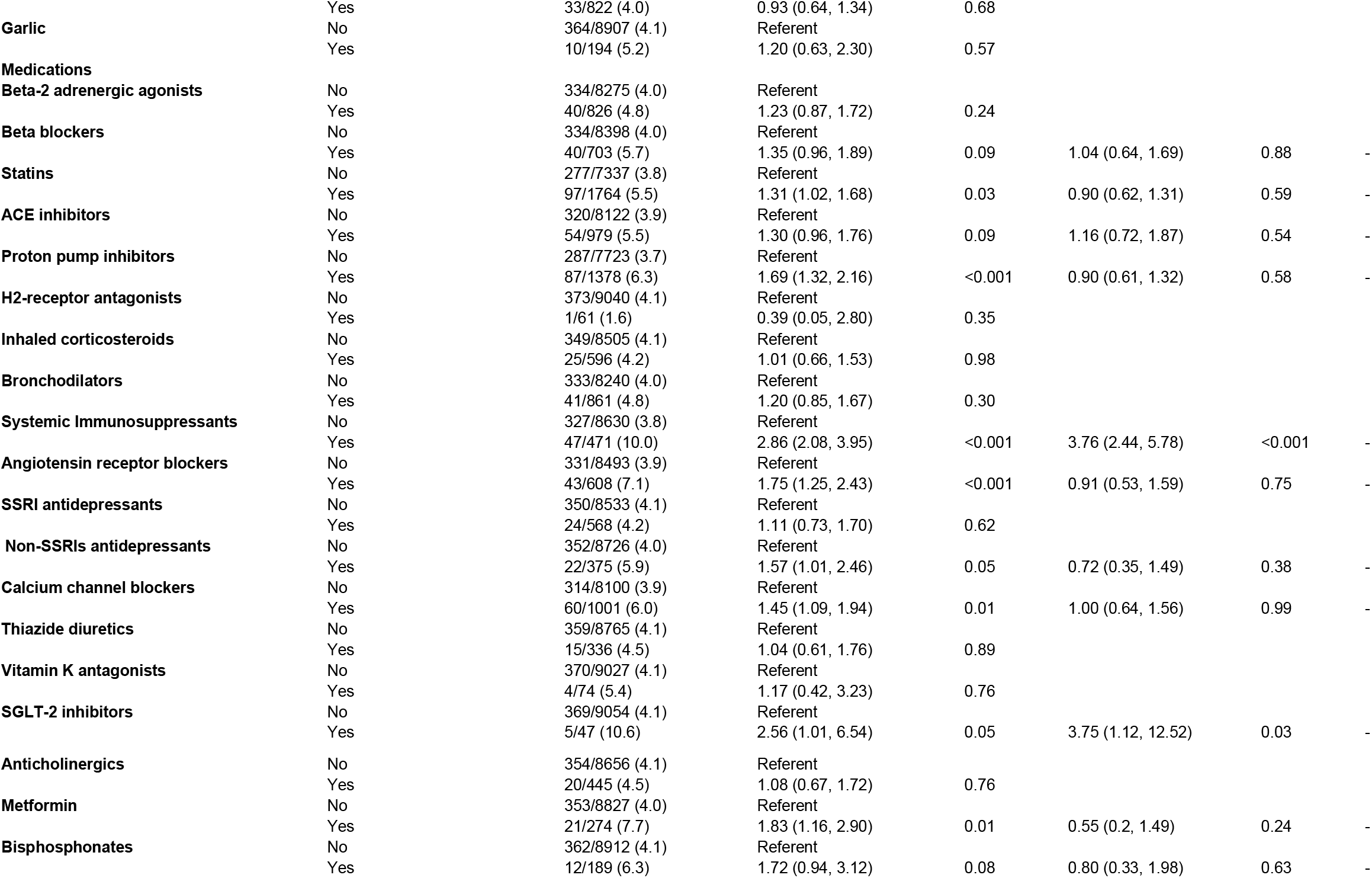
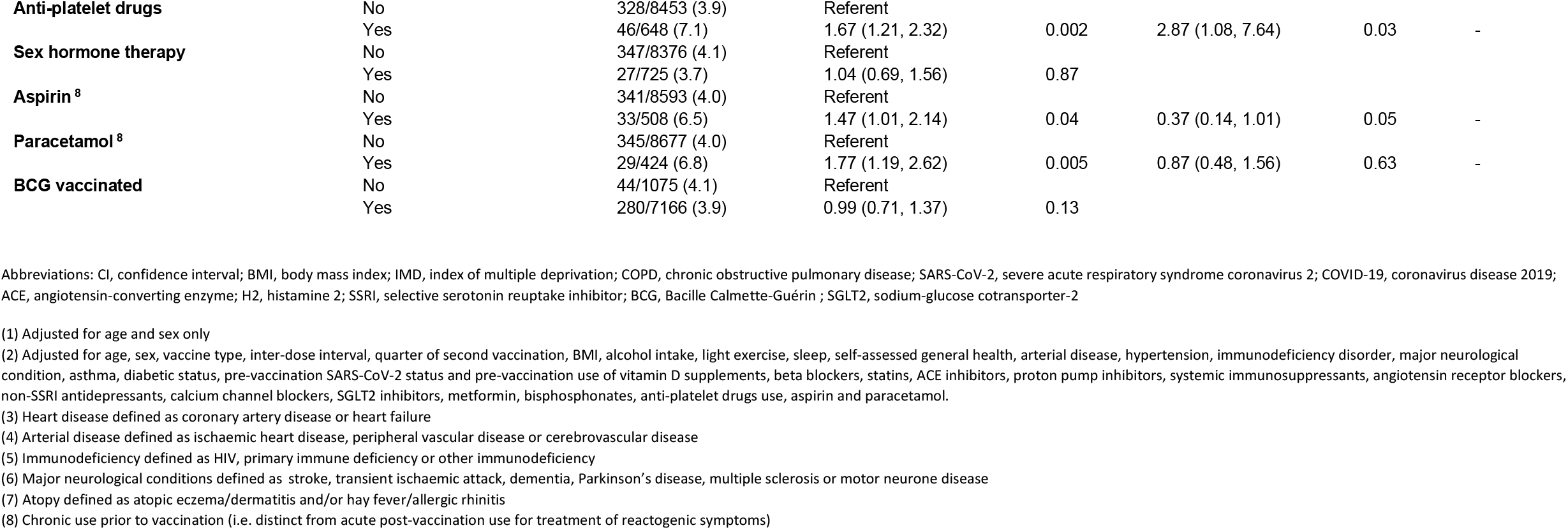
Determinants of post-vaccination seronegativity, all participants (n=9,101)

### Determinants of post-vaccination antibody titres in subset of individuals who were seropositive following a primary course of SARS-CoV-2 vaccination

To identify factors influencing the magnitude of antibody responses to COVID-19 vaccination, we investigated determinants of SARS-CoV-2 antibody titres in the subset of 8,727 participants who were seropositive following their second COVID-19 vaccine dose. Results of this analysis are presented in Table 3. A total of 22 factors associated with antibody titres after adjusting for age and sex with P<0.10 and were fitted in a fully adjusted model. Nine factors remained independently associated with antibody titres in the fully adjusted model with P<0.05. Six of these independently associated with lower antibody titres: receipt of ChAdOx1 *vs* BNT162b2 vaccine (43.4% lower titres, 95% CI 41.8-44.9, Figure 1); longer duration between date of second vaccine dose and date of sampling (6.8% lower, 2.3-11.1, for 5-8 weeks *vs* 2-4 weeks; 12.1% lower, 7.5-16.4, for 9-16 weeks *vs* 2-4 weeks, and 11.1% lower, 4.2-17.4, for >16 weeks *vs* 2-4 weeks); shorter interval between first and second vaccine doses (11.1% lower, 4.2-17.4, for <6 weeks vs >10 weeks; 5.8% lower, 3.3-8.3, for 6-10 weeks vs >10 weeks); receiving the second vaccine dose during the first quarter (Q1) or the final quarter (Q4) of the year (8.0% lower, 3.1-12.7, for Q1 *vs* Q2, and 47.7% lower, 11.4-69.1, for Q4 *vs* Q2); age ≥80 years (19.4% lower, 0.5-34.7 *vs* age <30 years), and presence of hypertension (4.0% lower, 1.0-6.9). Three factors independently associated with higher post-vaccination antibody titres: South Asian ethnicity (17.3% higher, 3.9-32.4, *vs* White ethnicity) or Mixed/Multiple/Other ethnicity (12.2% higher, 3.0-22.1, *vs* White ethnicity); higher body mass index (BMI; 2.9% higher, 0.2-5.7, for BMI 25-30 *vs* <25 kg/m^2^); and pre-vaccination seropositivity for SARS-CoV-2 (39.4% higher, 34.4-44.6, for individuals who were seropositive without COVID-19 symptoms pre-vaccination *vs* those who were seronegative pre-vaccination, and 117.9% higher, 105.6-130.9, for individuals who were seropositive with COVID-19 symptoms pre-vaccination *vs* those who were seronegative pre-vaccination). All the above determinants of post-vaccination antibody titres remained significant (P<0.05) when we performed a sensitivity analysis excluding 23 individuals who reported RT-PCR- or lateral flow test-confirmed SARS-CoV-2 infection after their second vaccine dose, but before their serology sampling date (data not shown).

**Table 3.**
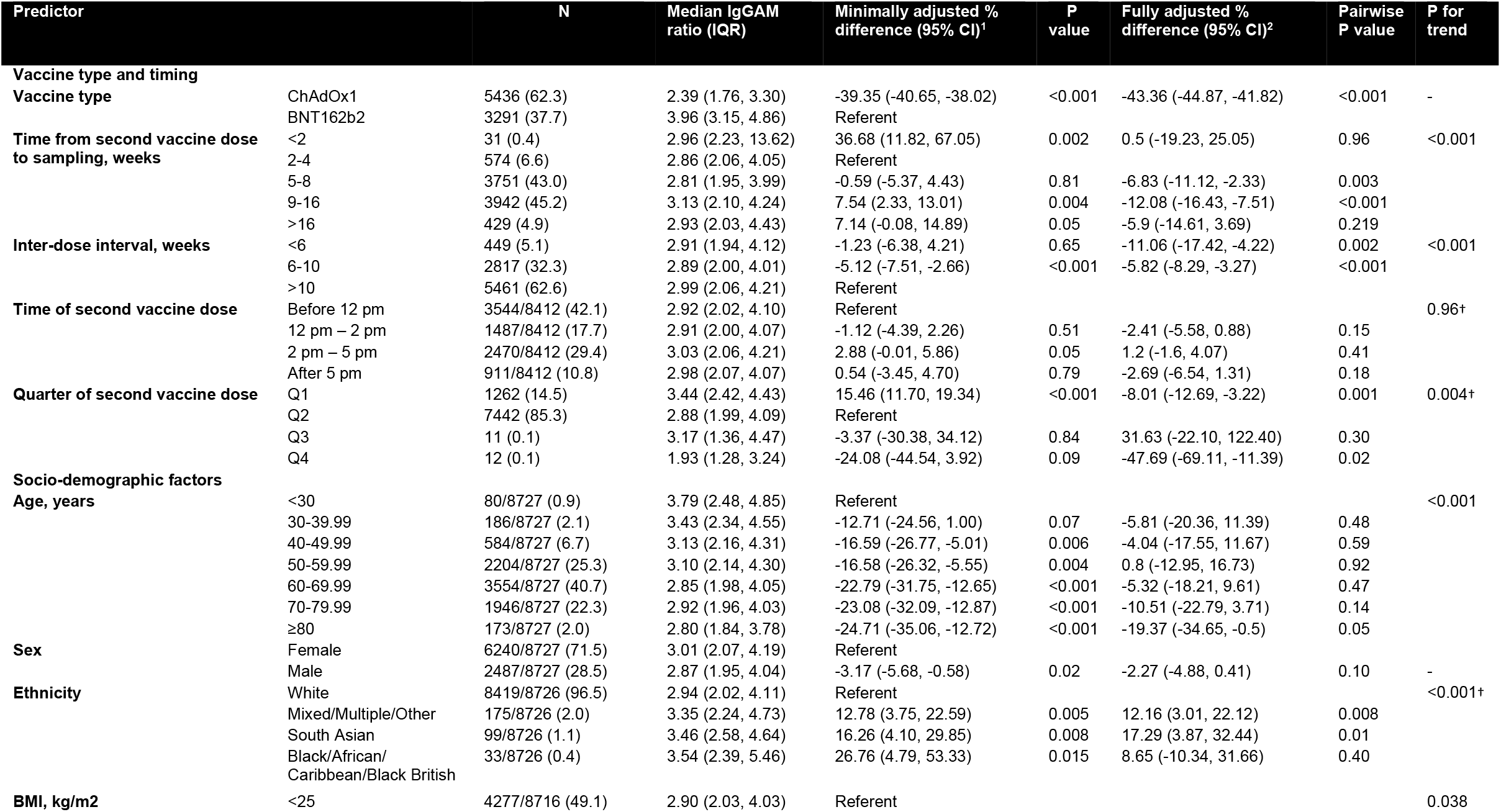
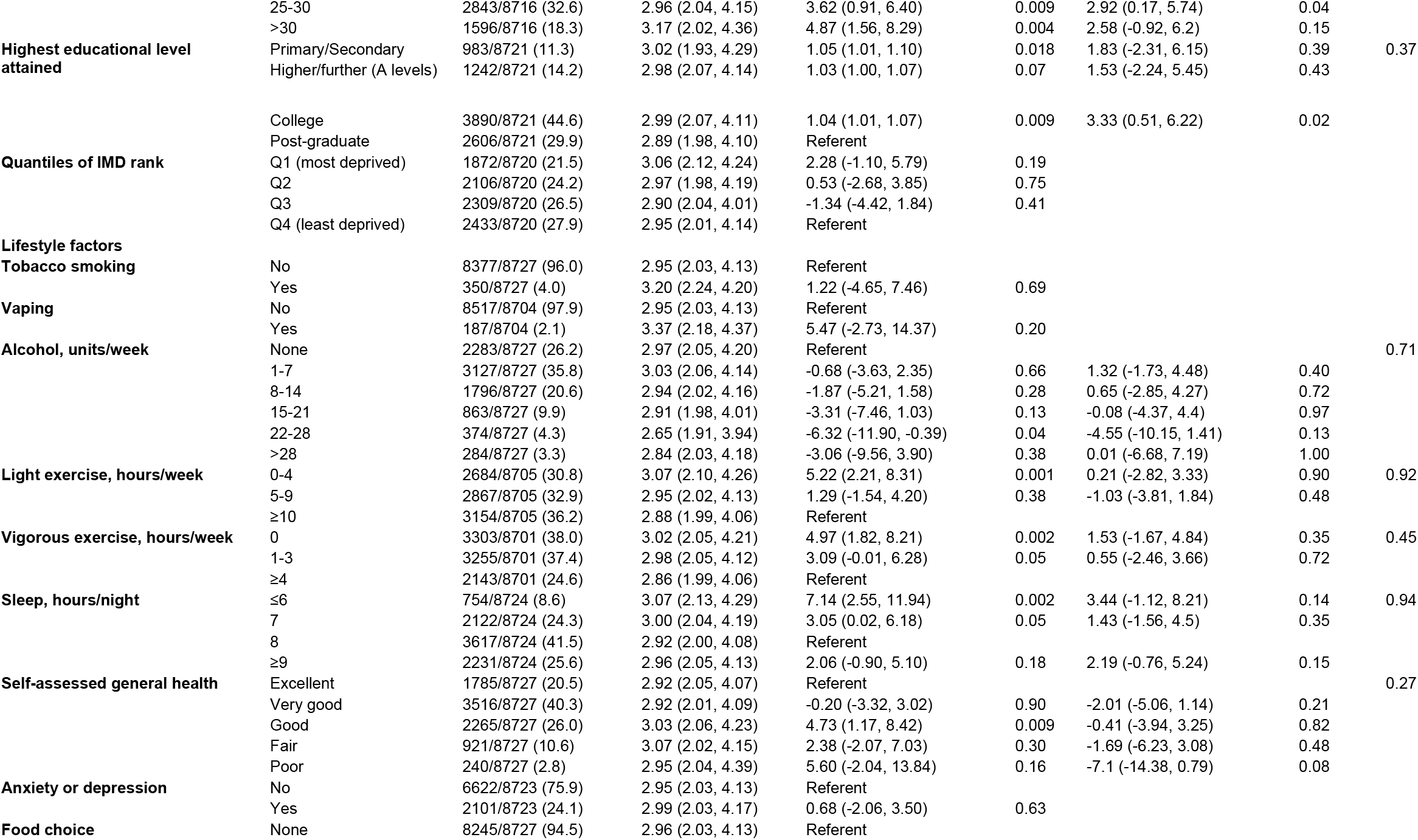
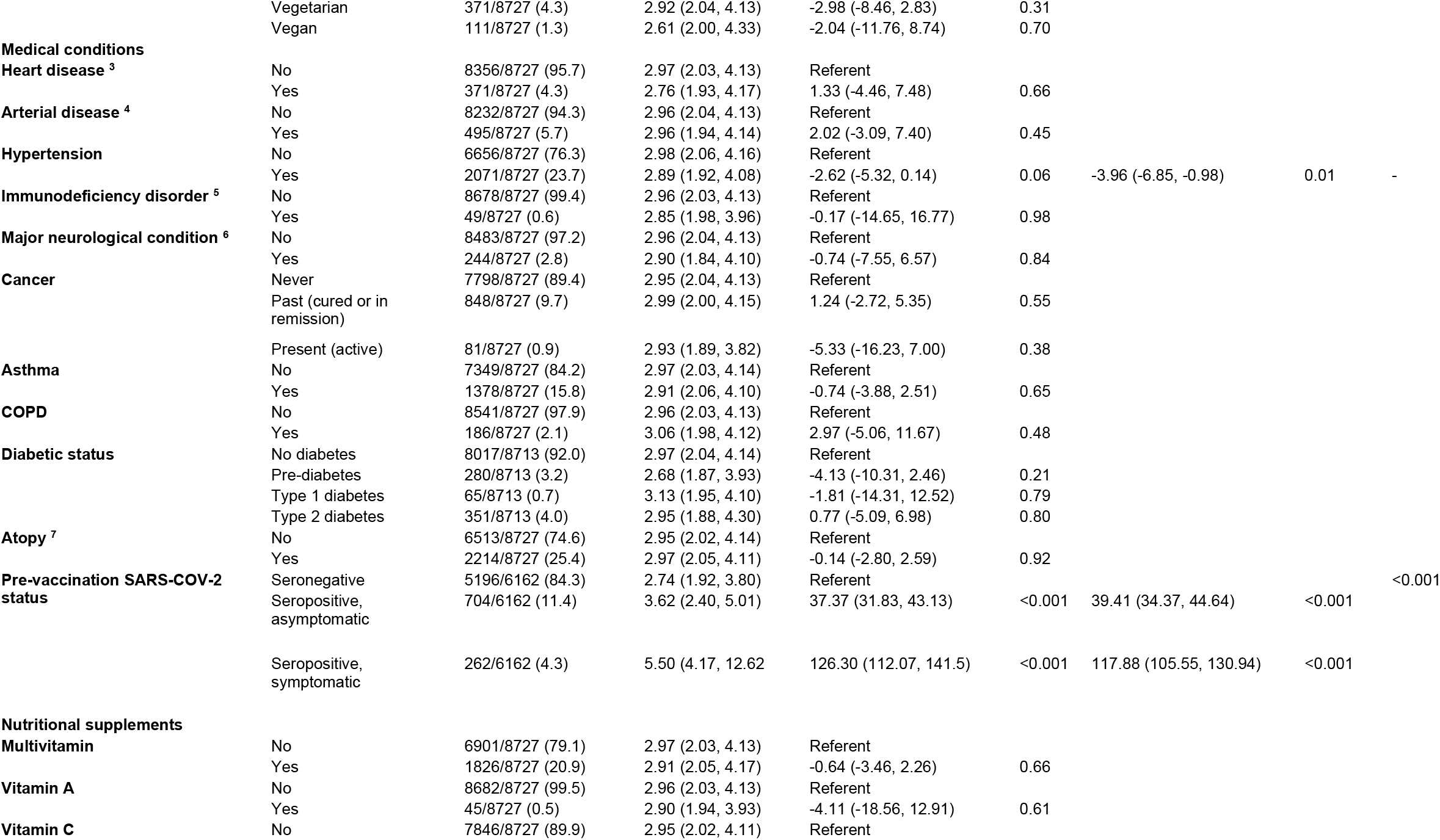
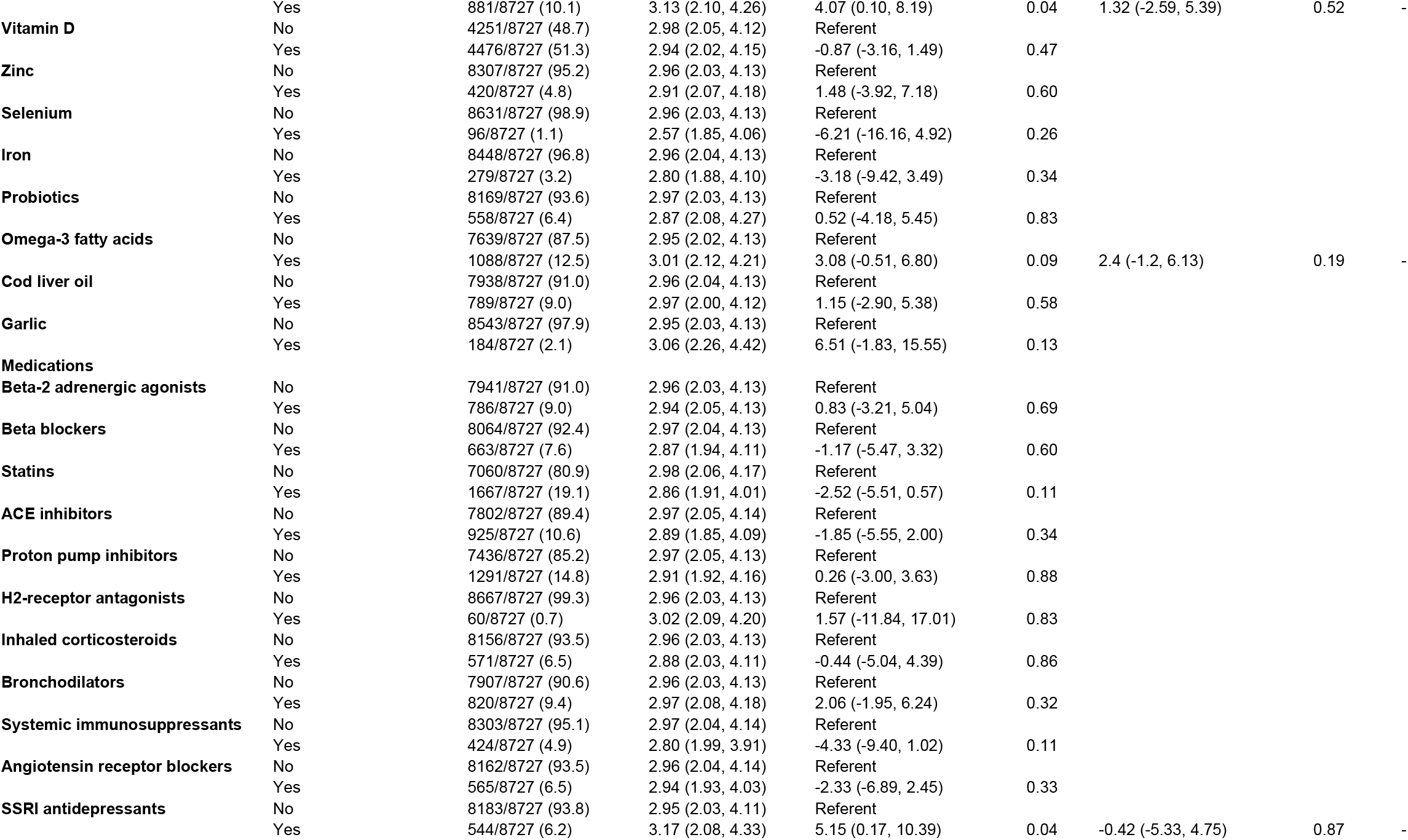
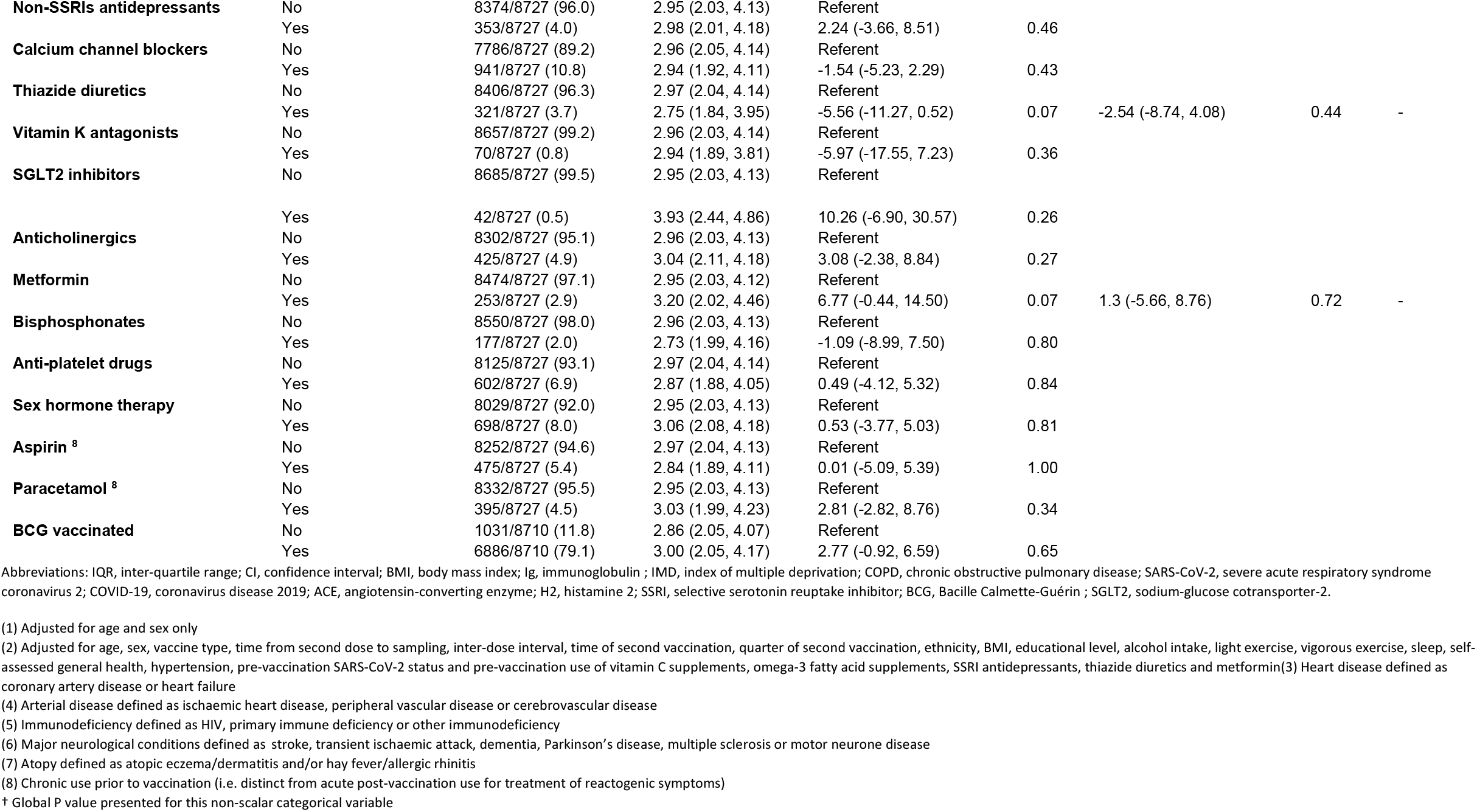
Determinants of post-vaccination antibody titres, subset of seropositive participants (n=8,727)

**Figure 1.**
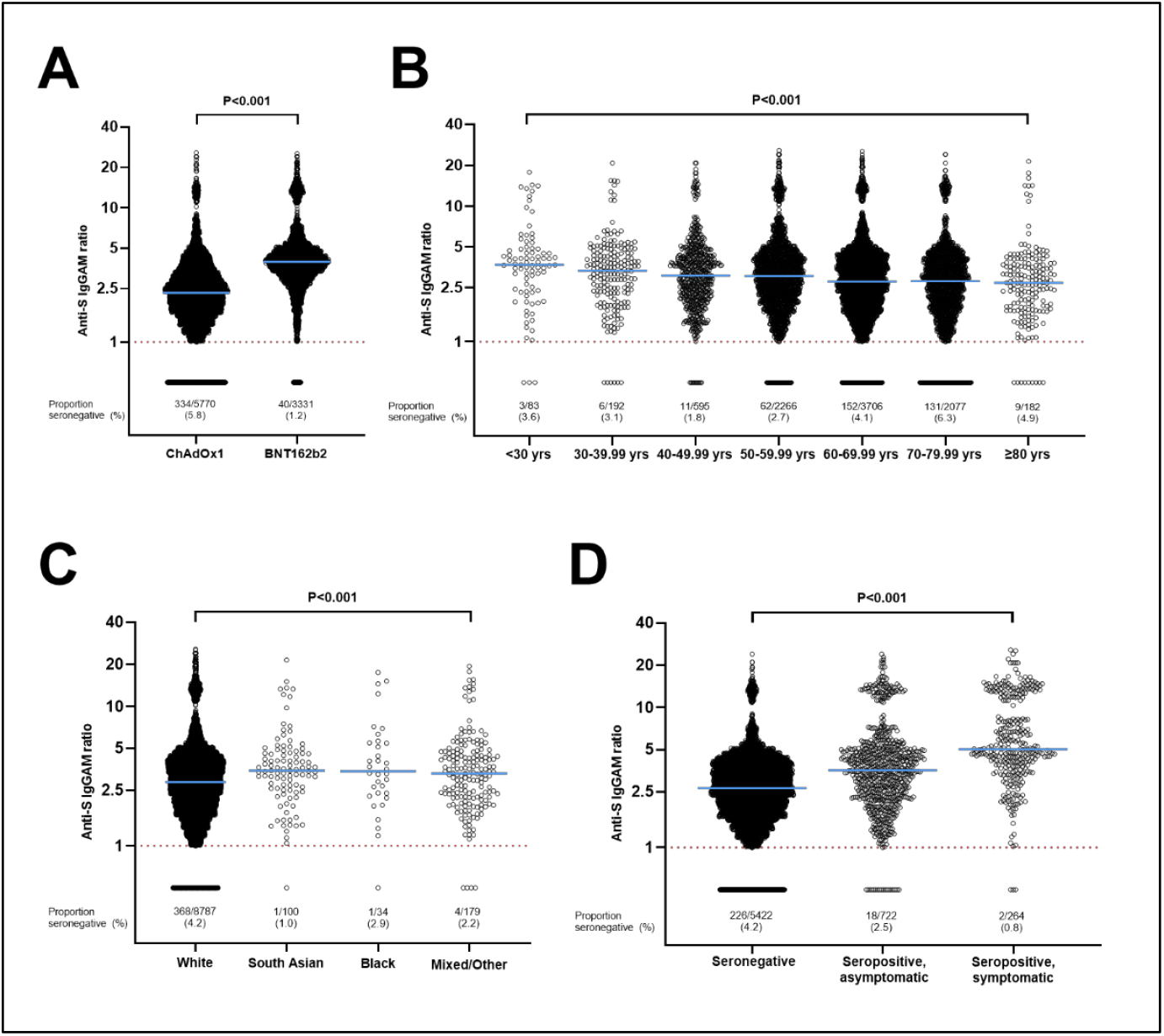
Antibody titres following two doses of vaccine by type of vaccine (A), age (B), ethnicity (C) and pre-vaccination SARS-CoV-2 status (D). For C, South Asian indicates people who self-identified their ethnic origin as Indian, Pakistani, or Bangladeshi, and Black indicates people who self-identified their ethnic origin as Black, African, Caribbean or Black British. P values from Mann Whitney test (A) and Kruskal Wallis tests (B-D). Anti-S, anti-spike; IgGAM, immunoglobulin G, A or M. Dotted line = limit of detection.

### Stratification of antibody responses by vaccine type

To explore whether determinants of antibody responses to vaccination were consistent for ChAdOx1 *vs* BNT162b2, we stratified the analysis of post-vaccination antibody titres according to type of vaccine received. Tables S4 and S5 (Supplementary Appendix) present the results of these analyses. Seven factors associated independently with post-vaccination antibody titres in ChAdOx1 recipients, but not in BNT162b2 recipients: lower titres in this group were associated with active cancer, consumption of selenium supplements and use of statins, Angiotensin Converting Enzyme (ACE) inhibitors and proton pump inhibitors (PPI), while South Asian and Mixed/Multiple/Other ethnic origin associated with higher titres. Conversely, three factors associated independently with lower post-vaccination antibody titres in BNT162b2 recipients, but not in ChAdOx1 recipients: older age, hypertension and use of systemic immunosuppressant medication. To test whether vaccine type modified the effects of these independent variables on post-vaccination antibody titres, we fitted all significant factors in our main multivariable model and included an interaction term for vaccine type. Three factors showed a small but statistically significant interaction: hypertension (P_interaction_=0.035), regular consumption of selenium supplements (P_interaction_=0.021) and use of systemic immunosuppressant medication (P_interaction_=0.046; Figure S2, Supplementary Appendix).

### Influence of post-vaccination paracetamol / NSAIDs on antibody response to primary course of vaccination

Results of an exploratory analysis to determine the influence of taking paracetamol or NSAIDs to treat post-vaccination symptoms are presented in Table S6. After fitting post-vaccination paracetamol/NSAID use into the main multivariable model we found a significant positive association between this factor and post-vaccination antibody titres. We then reasoned that this association could be confounded by an association between presence or severity of post-vaccination symptoms and higher post-vaccination antibody titres. After additionally correcting for report of post-vaccination symptoms (i.e., headache, fever, local soreness), the association between post-vaccination paracetamol/NSAID use and post-vaccination titres was rendered statistically non-significant.

### Antibody responses to booster doses in participants who were seronegative after two vaccine doses

Anti-spike Ig GAM titres for the subset of participants who provided dried blood spot samples following administration of a third vaccine dose are presented in Figure 2. Of 247 participants who had undetectable anti-spike IgGAM following two vaccine doses, all but eight (3.2%) had detectable antibodies following subsequent administration of a single booster dose. Details of the vaccine regimens, medical history and medication for the eight participants who remained seronegative following their booster dose are presented in Table S7, Supplementary Material: seven were taking immunosuppressant medication (five for autoimmune disease, two following solid organ transplant) and one had a primary antibody deficiency. Median post-booster antibody titres were higher among participants who were seropositive *vs* seronegative after two vaccine doses.

**Figure 2.**
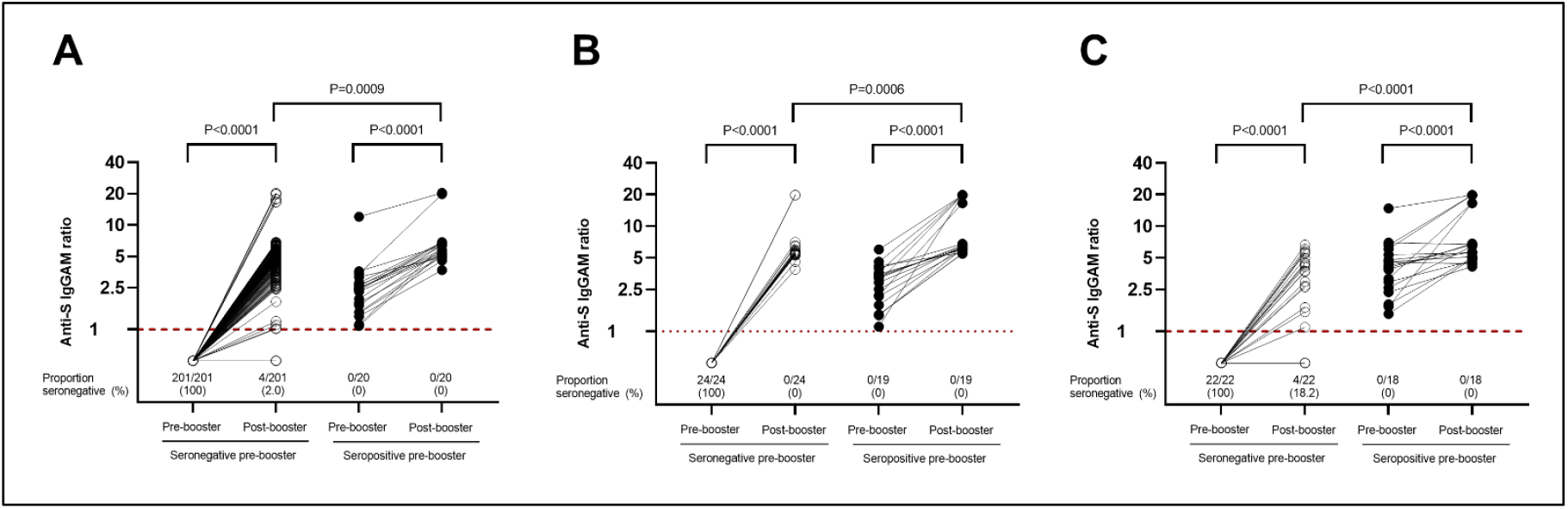
Anti-spike IgG/A/M titres before and after booster doses in participants who were seronegative *vs* seropositive after their primary course of SARS-CoV-2 vaccination. (A), participants receiving ChAdOx1 primary course with BNT162b2 booster. (B), participants receiving ChAdOx1 primary course with mRNA-1273 booster. (C), participants receiving BNT162b2 primary course with BNT162b2 booster. P values for paired / unpaired comparisons from Wilcoxon matched pairs signed rank tests and Mann Whitney tests, respectively. Anti-S, anti-spike; IgGAM, immunoglobulin G, A or M. Dotted line = limit of detection.

## Discussion

We present results of a large population-based study examining a comprehensive range of potential determinants of combined IgG, IgA and IgM responses to two very widely used SARS-CoV-2 vaccines. A major finding was that 5.8% of participants did not have detectable IgG/A/M anti-spike antibodies following two doses of ChAdOx1, as compared with 1.2% of those who had received two doses of BNT162b2 (aOR 7.03, 95% CI 4.39 to 11.24). Other risk factors for post-vaccination seronegativity included shorter inter-dose interval, older age, poorer general health, immunodeficiency disorder and use of systemic immunosuppressive medication. By contrast, pre-vaccination SARS-CoV-2 seropositivity and regular use of vitamin D supplements at the time of vaccination were associated with lower risk of post-vaccination seronegativity. Post-vaccination antibody titres were higher in participants of South Asian and mixed/other ethnic origin *vs* White participants, and in those who were overweight *vs* normal weight; these associations were independent of pre-vaccination SARS-CoV-2 serostatus. Post-vaccination use of paracetamol or NSAIDs was not associated with lower antibody responses to vaccination, after adjustment for post-vaccination symptoms, and determinants of post-vaccination anti-spike titres were not substantively different for participants who received two doses of ChAdOx1 *vs* BNT162b2. Single ‘booster’ doses of BNT162b2 or mRNA-1273 were highly effective in inducing seroconversion in the vast majority of adults who had undetectable anti-spike IgGAM antibodies after their first two doses of vaccine; those who failed to seroconvert after a booster dose were all taking immunosuppressant medication or had a primary immunodeficiency.

Our findings confirm previous reports of higher antibody responses to BNT162b2 *vs* ChAdOx1.^4,13,27^ Pre-existing immunity to adenovirus vectors may be one of the factors underlying this difference,^28^ and experiments to compare the prevalence of adenovirus neutralising antibody in pre-vaccination serum of participants who did *vs* did not develop anti-spike antibodies in response to ChAdOx1 are planned. Our finding that all participants who were seronegative after two doses of ChAdOx1 or BNT162b2 and who did not have an underlying immune defect seroconverted after receiving a single booster dose of BNT162b2 or mRNA-1273 provides strong support for roll-out of third vaccine doses to achieve near-universal seroconversion in the general population.

Our results with respect to the adverse influence of older age, shorter inter-dose interval, poorer general health, immunodeficiency and immunosuppressants on post-vaccination antibody responses are also broadly consistent with those of others^4-9^ – as is the finding that pre-vaccination SARS-CoV-2 infection associates with higher antibody titres after vaccination.^9,12,13^ However, we also report a number of novel findings. The independent association between regular use of vitamin D supplements and reduced risk of post-vaccination seronegativity echoes results of a previous study where we showed that high-dose vitamin D replacement enhances antigen-specific cellular responses following vaccination against varicella zoster virus;^29^ an intervention study nested within a randomised controlled trial of vitamin D supplementation for prevention of COVID-19 (ClinicalTrials.gov identifier NCT04579640) is currently on-going to explore this finding further. Associations between increased risk of post-vaccination seronegativity and use of sodium-glucose co-transporter-2 (SGLT2) inhibitors or anti-platelet drugs have also not previously been reported to our knowledge. Both drug classes are recognised to have immunomodulatory actions,^30,31^ but further studies are needed to determine whether the associations we report can be replicated in other cohorts.

We also found that White ethnicity and lower BMI both associated with lower post-vaccination antibody titres, independently of pre-vaccination SARS-CoV-2 serostatus. Ethnic variation in SARS-CoV-2 vaccine immunogenicity has not been widely studied to date: one study has reported lower antibody responses to vaccination in Jewish *vs* non-Jewish health care workers in Israel,^9^ while a population-based UK study reported higher antibody titres in ‘non-Whites’ *vs* Whites. However, pre-vaccination serostatus was not available for all participants in these studies; this omission is potentially important, because unvaccinated people of South Asian ethnic origin were at higher risk of SARS-CoV-2 infection in the pre-vaccination era.^20,24^ We demonstrate for the first time that higher anti-spike titres among vaccinated people of South Asian origin are not attributable to the higher rates of pre-vaccination SARS-CoV-2 infection that we have previously reported.^20,24^ The reasons for this phenomenon require further investigation: ethnic variation in recognition of SARS-CoV-2 T cell epitopes is recognised,^32^ and it may be that analogous variation in B cell epitopes underlies the ethnic differences in antibody responses to vaccination seen here. By contrast with ethnicity, several studies have investigated the impact of BMI on post-vaccination antibody titres. Their results are inconsistent, with one reporting lower antibody responses in those with BMI >30 kg/m^2^,^4^ another reporting higher responses in those with higher BMI^33^ and two reporting no association.^34,35^ Of note, studies investigating antibody responses to influenza vaccination report higher titres initially in obese participants, but a greater decline thereafter.^36^ This finding highlights the importance of longer term follow-up to elucidate the influence of BMI on humoral responses to vaccination.

We also report some important null results. The lack of association between time of day of vaccination and degree of antibody response conflicts with findings of studies which variously report higher anti-spike responses in those vaccinated earlier^37^ or later ^38^ in the day. We also show no association between pre- or post-vaccination use of paracetamol or NSAIDs and antibody responses, after adjustment for incidence of post-vaccination symptoms. This provides reassurance that use of these medications to manage reactive post-vaccination symptoms does not compromise SARS-CoV-2 vaccine immunogenicity. We also show no association between higher titres and socioeconomic deprivation, as reported by others; ^4,7^ this may reflect the fact that other studies did not adjust for pre-vaccination serostatus, which is more likely to be positive in socioeconomically deprived populations. There was no evidence of association for lifestyle factors hypothesised to influence vaccine responses including dietary factors, use of micronutrient supplements other than vitamin D, sleep, alcohol use and tobacco smoking.

Our study has several strengths. We utilised a CE-marked assay with high sensitivity and specificity that targets three different types of antibody,^21^ and that we have validated as a correlate of protection against breakthrough disease.^22,23^ Its large size affords power to detect determinants of having undetectable anti-spike antibodies after SARS-CoV-2 vaccination – a relatively rare, but potentially clinically important outcome. The population-based nature of this study, and the fact that we investigated effects of two major vaccines utilising differing technology (one live vector, one mRNA) enhances the generalisability of our findings. Very detailed characterisation of participants allowed us to investigate a wide range of potential determinants of vaccine immunogenicity, and to adjust for multiple confounders, including pre-vaccination serostatus.

Our study also has some limitations. We did not study cellular responses to vaccination, and these may be discordant with humoral responses.^10,27^ We also studied responses at an early time point only; a high early peak in antibody titres following vaccination is not necessarily an indicator of durable response.^36^ As with any observational study, we cannot exclude the possibility that some of the associations we report might be explained by residual or unmeasured confounding. However, we have minimised the risk of this by adjusting for a comprehensive list of putative determinants of vaccine immunogenicity. COVIDENCE UK is also a self-selected cohort, and thus certain demographics—such as people <30 years old, people of lower socioeconomic status, and non-White ethnic groups—are under-represented. This may have limited our power to investigate antibody responses in some sub-groups, such as participants of Black ethnicity (who are at higher risk of SARS-CoV-2 infection^39-41^ and adverse outcomes^42^ than White people).

In conclusion, this large population-based study reports on the influence of a comprehensive range of potential sociodemographic, behavioural, clinical, pharmacological and nutritional determinants on antibody responses to two major SARS-CoV-2 vaccines. Importantly, we also show that booster doses of mRNA vaccines are highly effective in achieving seroconversion in healthy people who fail to mount antibody responses to SARS-CoV-2 trimeric spike glycoprotein after receiving two doses of ChAdOx1.

## Supporting information

Supplementary Appendix Jolliffe et al

## Data Availability

De-identified participant data will be made available upon reasonable request to the corresponding author, subject to terms of research ethics committee approval and Sponsor requirements.

## Contributors

ARM wrote the study protocol, with input from DAJ, MT, and SOS. HH, MT, JS, GAD, RAL, CJG, FK, AS, and ARM contributed to questionnaire development and design. DAJ co-ordinated and managed the study, with input from ARM, NP, HH and SM. HH, JS, ARM, and SOS supported recruitment. SEF and AGR developed, validated, and performed laboratory assays. DAJ, MT, HH, MG, GV and FT contributed to data management. DAJ and ARM directly accessed and verified the data. Statistical analyses were done by DAJ, with input from ARM. DAJ and ARM wrote the first draft of the report. All authors revised it critically for important intellectual content, gave final approval of the version to be published, and agreed to be accountable for all aspects of the work in ensuring that questions related to the accuracy or integrity of any part of the work were appropriately investigated and resolved. DAJ, MT, HH, GV and FT had full access to all data in the study, and ARM had final responsibility for the decision to submit for publication.

## Data sharing

De-identified participant data will be made available upon reasonable request to the corresponding author, subject to terms of Research Ethics Committee approval and Sponsor requirements.

## Acknowledgments

This study was supported by a grant from Barts Charity to ARM and CJG (MGU0466) and by donations to Queen Mary University of London from the Fischer Family Trust, the Exilarch’s Foundation and DSM Nutritional Products Ltd. DAJ is supported by a Barts Charity Lectureship (MGU0459). MT is supported by a grant from the Rosetrees Trust and The Bloom Foundation (M771). The work was carried out with the support of BREATHE - The Health Data Research Hub for Respiratory Health (MC_PC_19004) in partnership with SAIL Databank. BREATHE is funded through the UK Research and Innovation Industrial Strategy Challenge Fund and delivered through Health Data Research UK. The views expressed are those of the authors and not necessarily those of the funders. We thank all participants of COVIDENCE UK, and the following organisations who supported study recruitment: Asthma UK, the British Heart Foundation, the British Lung Foundation, the British Obesity Society, Cancer Research UK, Diabetes UK, Future Publishing, Kidney Care UK, Kidney Wales, Mumsnet, the National Kidney Federation, the National Rheumatoid Arthritis Society, the North West London Health Research Register (DISCOVER), Primary Immunodeficiency UK, the Race Equality Foundation, SWM Health, the Terence Higgins Trust, and Vasculitis UK.

