## Supplementary Appendix Jolliffe et al for "Determinants of antibody responses to two doses of ChAdOx1 nCoV-19 or BNT162b2 and a subsequent booster dose of BNT162b2 or mRNA-1273: population-based cohort study (COVIDENCE UK)"

**Supplementary Methods**

### Sample size

Using the Stata powerlog program, we estimated that a minimum sample size of 8,192 would be required to detect a difference of at least 2% in the proportion of exposed *vs* unexposed participants experiencing a given binary outcome (equivalent to an adds ratio [OR] of 1.08), with 80% power, for a binary exposure with maximum variability (probability 0.50 changing to 0.52) and a moderate correlation ($R^2$=0.4) with other variables in a logistic regression model, using a two-sided test and 5% significance. The post-vaccination antibody study was a pragmatic study including all participants meeting the inclusion criteria, with no sample size specified.

### Statistical analysis

Statistical analyses were performed using Stata (version 14.2). Logistic regression models were used to estimate adjusted ORs, 95% CIs and associated pairwise P-values for potential determinants of post-vaccination seronegativity in all participants. Linear regression models were used to estimate beta-coefficients, 95% CIs and associated pairwise P-values for potential determinants of log-transformed antibody titres in the subset of seropositive participants. For ease of interpretation, log-transformed estimates of antibody titres were exponentiated and a percentage increase or decrease was calculated for every one-unit increase in the potential determinant. We first estimated ORs and beta-coefficients in minimally adjusted models, and included factors independently associated with each outcome at the 10% significance level in fully adjusted models. Both the minimally adjusted and fully adjusted models were controlled for age (<30 years, 30 to <40 years, 40 to <50 years, 50 to <60 years, 60 to <70 years, 70 to <80 years and ≥80 years) and sex (male *vs* female). For fully adjusted models, we additionally calculated a P-value for trend for scalar independent variables with more than 2 levels, or a global P-value for non-scalar independent variables with more than 2 levels. Pairwise associations having P-values <0.05 were not considered significant in fully adjusted models where the global P-value or P-value for trend for the independent variable in question was ≥0.05. Paired comparisons of antibody titres were conducted using Wilcoxon matched pairs signed rank tests and unpaired comparisons were conducted using Mann-Whitney U tests (for 2 groups) and Kruskal Wallis tests (for three or more groups). Analyses were done for all participants with available data only; missing data were not imputed. Correction for multiple comparisons was not applied, on the grounds that we were testing *a priori* hypotheses for all independent variables investigated.[1]

In a sensitivity analysis, we excluded participants from the analysis of antibody titres after two vaccine doses who reported a positive RT-PCR or lateral flow test for SARS-CoV-2 between the date of their second dose of vaccine and the date on which they provided their dried blood spot sample. We also stratified the analysis of antibody titres following two vaccine doses according to the type of vaccine received to explore whether determinants of antibody responses to two vaccine doses were consistent for ChAdOx1 *vs* BNT162b2. Given reports that peri-vaccination use of antipyretic analgesics may attenuate vaccine immunogenicity,[2] we also conducted an exploratory analysis to determine the influence of taking paracetamol or non-steroidal anti-inflammatory drugs (NSAIDs) to treat post-vaccination symptoms on post-vaccination antibody titres.

**Table S1.** Baseline questions

| **Sociodemographic** | |
|---|---|
| Date of birth (DD/MM/YYYY) | |
| Post code | |
| Address | |
| Please state your **assigned sex at birth.** | • Male<br>• Female |
| What is your **current** height? (if you are unsure, please put your best estimate) | -Feet/inches<br>-Centimetres |
| What is your **current** weight? | - Stones (sts) / pounds (lbs)<br>- Kilograms (kg) |
| What is your ethnic origin? | • White<br>▪ English / Welsh / Scottish / Northern Irish / British<br>▪ Irish<br>▪ Gypsy or Irish Traveller<br>▪ Any other white background<br>• Mixed / Multiple ethnic groups<br>▪ White and Black Caribbean<br>▪ White and Black African<br>▪ White and Asian<br>▪ Any other Mixed / Multiple ethnic backgrounds<br>• Asian / Asian British<br>▪ Indian<br>▪ Pakistani<br>▪ Bangladeshi<br>▪ Chinese<br>▪ Any other Asian background<br>• Black / African / Caribbean / Black British<br>▪ African<br>▪ Caribbean<br>▪ Any other Black / African / Caribbean background<br>• Arab<br>• Other Ethnic Group |
| What is the highest level of education that you have completed? | • Primary school (0)<br>• Secondary school up to 16 years (1)<br>• Higher or secondary or further education (A-levels, BTEC, etc.) (2)<br>• College or university (3)<br>• Post-graduate degree (4) |
| Which of the following best describes your current occupational status? | • Employed<br>• Self-employed<br>• Retired<br>• Furloughed<br>• Unemployed<br>• Student<br>• Other |

| Comorbidities | |
|---|---|
| Have you ever been diagnosed with any of the following conditions by a doctor?<br><br>Select all that apply. | • Asthma<br>• Atopic Eczema or Atopic Dermatitis<br>• Autoimmune disease (e.g. rheumatoid arthritis, multiple sclerosis (MS), lupus (SLE), Crohn's disease, ulcerative colitis, psoriasis, Raynaud's disease, scleroderma)<br>• Cancer<br>• Cerebral Palsy<br>• COPD (including chronic bronchitis, and emphysema)<br>• Cystic Fibrosis<br>• Dementia<br>• Diabetes or pre-diabetes<br>• Hayfever or Allergic Rhinitis<br>• Heart Attack, Angina or Coronary Artery Disease<br>• Heart Failure<br>• High Blood Pressure (Hypertension)<br>• HIV Infection<br>• Hyperparathyroidism (overactive parathyroid gland)<br>• Kidney stones<br>• Other kidney disease<br>• Leg Artery Disease (also known as 'peripheral vascular disease', 'peripheral arterial disease' or 'intermittent claudication')<br>• Mental health disorder<br>• Motor Neurone Disease<br>• Organ transplant<br>• Parkinson's Disease<br>• Primary immune deficiency (e.g. antibody deficiency, combined immunodeficiency)<br>• Sarcoidosis<br>• Sickle Cell Disease (i.e. two copies of altered gene, affected by anaemia and other complications<br>• Sickle Cell Carrier (also known as 'sickle cell trait', with only one copy of altered gene: few symptoms if any)<br>• Splenectomy (removal of spleen)<br>• Stroke or Mini-Stroke<br>• Tuberculosis (TB)<br>• None of the above |
| You indicated you have been diagnosed with diabetes or pre-diabetes. Please specify your diagnosis: | • Pre-diabetes (high blood sugar levels, not enough to be diagnosed with diabetes)<br>• Type 1 diabetes<br>• Type 2 diabetes<br>• Other type of diabetes |
| Do you currently have cancer? | • Never<br>• No, cancer cured or in remission<br>• Yes, currently receiving treatment |
| Under each heading, please click the ONE box that best describes your health TODAY.<br>(Anxiety / Depression) | • I am not anxious or depressed<br>• I am moderately anxious or depressed<br>• I am extremely anxious or depressed |
| Over the last 12 months, would you say that on the whole, your health has been: | • Excellent<br>• Very good<br>• Good<br>• Fair |

| | |
|---|---|
| | • Poor |
| **Vaccination** | |
| Have you ever had the BCG vaccine?<br><br>*This is the vaccine against Tuberculosis (TB), it's injected in the upper arm and usually leaves a small scar* | • Yes<br>• No<br>• Unsure |
| **Lifestyle** | |
| Which of these best describes your use of cigarettes? | • I have never smoked cigarettes<br>• I used to smoke cigarettes occasionally but now not at all<br>• I used to smoke cigarettes daily but now not at all<br>• I smoke cigarettes occasionally but not every day<br>• I smoke cigarettes daily |
| Which of these best describes your use of e-cigarettes (vaping)? | • I have never vaped or used e-cigarettes<br>• I used to use e-cigarettes occasionally, but now not at all<br>• I used to use e-cigarettes daily but now not at all<br>• I vape occasionally but not every day<br>• I vape daily |
| During the last week, roughly how many hours did you spend doing light exercise that does not make you particularly breathless, such as light gardening, walking, including walking for pleasure or exercise, walking to the shops, walking to work? | - 0 -10+ hours |
| During the last week, roughly how many hours did you spend doing more vigorous physical exercise of sufficient intensity to make you breathless or to raise your heart rate significantly, such as heavy physical work, more strenuous gardening (e.g. vigorous digging, landscaping) swimming, jogging, aerobics, football, tennis, cycling, gym workout? | - 0 -10+ hours |
| During the past month, how many hours of actual sleep did you get per night on average?<br>(This may be different than the number of hours you spent in bed) | • 0–24 hours |
| **Diet** | |
| How many units of alcohol did you drink over the last 7 days?<br>*One unit is ½ a pint (285 ml) of ordinary beer, lager or cider; 25ml of spirits; 1 small glass (75ml) of wine; or 50ml of sherry* | • None<br>• 1–7 units<br>• 8–14 units<br>• 15–21 units<br>• 22–28 units<br>• More than 28 units |
| Do you **exclude** any of the following foods from your diet? Select all that apply. | - Eggs<br>- Cow's milk or products made from cow's milk (e.g. cheese, yoghurt)<br>- Fish<br>- White meat (e.g. poultry)<br>- Red meat<br>- No, I eat all of these foods |

| | |
|---|---|
| Over the **last month**, have you taken any of the following supplements at least **once per week**?<br><br>Select all that apply. | • Multivitamin (including prenatal multivitamins)<br>• Supplement containing vitamin A only<br>• Supplement containing vitamin B only<br>• Supplement containing vitamin C only<br>• Supplement containing vitamin D only<br>• Supplement containing calcium only<br>• Supplement containing calcium and vitamin D combined<br>• Supplement containing vitamin E only<br>• Supplement containing zinc only<br>• Supplement containing iron only<br>• Supplement containing probiotics<br>• Supplement containing fish oil, krill oil or other source of omega-3 fatty acids<br>• Supplement containing cod liver oil<br>• Supplement containing echinacea<br>• Supplement containing garlic or garlic powder (allicin)<br>• Supplement containing turmeric / curcumin<br>• Supplement containing Cannabidiol (CBD) oil<br>• Supplements containing folic acid<br>• Supplement containing Selenium only<br>• Other (e.g. other micronutrients (such as herbal supplements) or combinations of micronutrients (such vitamin C & zinc)) Please specify:<br>• None of the above |

**Medications**

Please type the names of all the medications you are currently taking below, one medication per box. The next pages will collect details about dosage for each.

Include all types of medications taken at home or administered in a hospital or clinic (capsules, tablets, contraceptive pills or implants, inhalers, injections, intravenous infusions, monoclonal antibodies, chemotherapy, immunosuppressants, etc.)

Please note that other pages will collect details about the amount of medicine in each dose (next page) and how often you take each dose (the page after that). If there are any details about your medication that aren't captured by our form (e.g. if you take different doses of a medicine at different times of day), there will be space to enter them in a blank text box at the end of this section of the questionnaire.

| | **Medication name (generic or brand name, either is fine)** |
|---|---|
| Medication 1 | |
| Medication 2 | |
| …. | |

**Table S2.** Monthly follow-up questions

| **Questions** | |
|---|---|
| Since you last checked in with us, have you had a nose or throat swab for COVID-19 or any other respiratory virus, or has a result from a previous swab test become newly available?<br>(This question is about tests to detect the virus itself: they are usually done in somebody who has symptoms, but screening of asymptomatic people can also be done. It's usually a nose/throat swab, but saliva tests are also becoming available) | • Yes<br>• No |
| On what date did you have this nose / throat swab?<br>If you are not sure of the exact date, enter the approximate date (DD/MM/YYYY). | |
| What was the result? Click as many as apply. | • Positive for COVID-19 (SARS-CoV-2 coronavirus)<br>• Positive for influenza virus<br>• Positive for another respiratory virus<br>• Negative for all/any viruses tested<br>• Not known |
| Since you last checked in with us, have you had an ANTIBODY test for COVID-19? (This is a test to detect whether or not you have already had the virus, usually done in somebody who does not currently have symptoms. It's usually a blood test / fingerprick test, but saliva tests are also available) | • Yes<br>• No |
| On what date did you have this ANTIBODY test? If you are not sure of the exact date, enter the approximate date (DD/MM/YYYY) | |
| What was the result? | • Positive<br>• Negative<br>• Not known |
| Since you last checked in with us, have you had a vaccine (immunisation) against COVID-19? | • Yes<br>• No |
| On what date did you have this COVID-19 vaccine (immunisation)? If you are not sure of the exact date, enter the approximate date (DD/MM/YYYY) | |
| Since you last checked in with us (last questionnaire sent <previous monthly questionnaire date>), have you had one or more doses of a COVID-19 vaccine (immunisation)? First doses and booster doses both count. | • Yes, I have had one or more doses of COVID-19 vaccine <strong>since I filled my last questionnaire<br>• No, I have not had any doses of COVID-19 vaccine <strong>since I filled my last questionnaire<br>• Not sure (e.g. you took part in vaccine trial, but dont yet know whether or not you had the real vaccine or the placebo (dummy vaccine) |
| How many doses of COVID-19 vaccine have you had since you last checked in with us? (last questionnaire sent 18th November 2021) | • One dose since my last questionnaire<br>• Two doses since my last questionnaire |
| On what date did you have the first of these vaccine doses? If you are not sure of the exact date, enter the approximate date (DD/MM/YYYY) e.g. for 16th December 2020, write 16/12/2020 | |

| | |
|---|---|
| On what date did you have the second of these vaccine doses? If you are not sure of the exact date, enter the approximate date (DD/MM/YYYY) e.g. for 16th December 2020, write 16/12/2020 | |
| Which COVID-19 vaccine did you have? | • Oxford / AstraZeneca / ChAdOx1<br>• Pfizer / BioNTech<br>• Moderna<br>• Valneva<br>• Novavax<br>• Janssen (also known as Johnson & Johnson)<br>• Other - please specify<br>• Not sure / don't know |
| Did you experience any of the following symptoms following your COVID-19 vaccine? | • Tenderness, soreness, swelling, redness or a painful, heavy feeling at the injection site<br>• Feeling tired<br>• Headache<br>• Fever / high temperature (37.8° C or greater)<br>• Muscle aches<br>• Swelling of the glands in your armpit or neck<br>• Other symptoms: specify<br>• None of the above |
| Did you take any of the following to treat post-vaccination symptoms (e.g. fever, muscle aches etc)? | • Paracetamol (e.g. Disprol, Hedex, Medinol, Panadol)<br>• Ibuprofen (e.g. Nurofen, Brufen, Calprofen)<br>• Something else: please write name here<br>• None of the above |
| Since you last checked in with us, have you experienced any of the following symptoms: cold or flu symptoms, sore throat, persistent cough, loss of smell or taste, fever, fatigue, diarrhoea, abdominal pain or loss of appetite? | • Yes, I have had one or more of these symptoms since completing my last COVIDENCE UK questionnaire<br>• No, I have not had any of these symptoms since completing my last COVIDENCE UK questionnaire |
| When did your symptoms start? (DD/MM/YYYY) | |
| Did you have a persistent cough (coughing a lot for more than an hour, or 3 or more coughing episodes in 24 hours)? | • No<br>• Persistent dry cough (i.e. producing little or no phlegm)<br>• Persistent productive cough |
| Did you experience unusual fatigue? | • No<br>• Mild fatigue<br>Severe fatigue – I struggled to get out of bed |
| Did you have a loss of sense of smell or taste? | • Yes<br>• No |
| Did you skip any meals because you felt unwell? | • Yes<br>• No |

**Table S3.** Participant characteristics, post-booster sub-study (n=304)

| Characteristic | | Seronegative pre-booster (n=247) | Seropositive pre-booster (n=57) | Overall (n=304) |
|---|---|---|---|---|
| Age | Median age, years (IQR) | 67.0 (61.9-72.6) | 65.9 (61.1, 69.6) | 66.6 (61.8-72.3) |
| | Age range, years | 29.5-87.2 | 42.4-85.3 | 29.5-87.2 |
| Sex, N (%) | Male | 92 (37.2) | 14 (24.6) | 106 (34.9) |
| | Female | 155 (62.8) | 43 (75.4) | (198 (65.1) |
| Ethnicity, N (%) [1] | White | 244 (98.8) | 55 (96.5) | 299 (98.4) |
| | South Asian | 1 (0.4) | 0 (0.0) | 1 (0.3) |
| | Black/African/Caribbean/Black British | 0 (0.0) | 0 (0.0) | 0 (0.0) |
| | Mixed/Multiple/Other | 2 (0.8) | 2 (3.5) | 1 (0.3) |
| Nation of residence [2] | England | 219 (86.7) | 52 (91.2) | 271 (89.1) |
| | Northern Ireland | 2 (0.8) | 0 (0.0) | 2 (0.7) |
| | Scotland | 18 (7.3) | 4 (7.0) | 22 (7.2) |
| | Wales | 8 (3.2) | 1 (1.8) | 9 (3.0) |
| Body mass index, kg/m$^2$, N (%) [3] | <25 | 114 (46.2) | 26 (45.6) | 140 (46.1) |
| | 25-30 | 88 (35.6) | 19 (33.3) | 107 (35.2) |
| | >30 | 45 (18.2) | 12 (21.1) | 57 (18.7) |
| Highest educational level attained, N (%) [4] | Primary/Secondary | 32 (13.0) | 5 (8.8) | 37 (12.2) |
| | Higher/Further (A levels) | 39 (15.8) | 7 (12.3) | 46 (15.1) |
| | College | 102 (41.2) | 26 (45.6) | 128 (42.1) |
| | Post-graduate | 74 (30.0) | 19 (33.3) | 93 (30.6) |
| Quantiles of IMD rank, N (%) [5] | Q1 (most deprived) | 68 (27.5) | 8 (14.0) | 76 (25.0) |
| | Q2 | 56 (22.7) | 20 (35.1) | 76 (25.0) |
| | Q3 | 60 (24.3) | 16 (28.1) | 76 (25.0) |
| | Q4 (least deprived) | 63 (25.5) | 13 (22.8) | 76 (25.0) |
| Tobacco smoking, N (%) | Non-current/never smoker | 240 (97.2) | 54 (94.7) | 294 (96.7) |
| | Current smoker | 7 (2.8) | 3 (5.3) | 10 (3.3) |
| Alcohol consumption/week, units, N (%) | None | 75 (30.4) | 18 (31.6) | 93 (30.6) |
| | 1-7 | 73 (29.6) | 24 (42.1) | 97 (31.9) |
| | 8-14 | 52 (21.1) | 7 (12.3) | 59 (19.4) |
| | 15-21 | 25 (10.1) | 6 (10.5) | 31 (10.2) |
| | 22-28 | 11 (4.5) | 0 (0.0) | 11 (3.6) |
| | >28 | 11 (4.5) | 2 (3.5) | 13 (4.3) |
| Self-assessed general health | Excellent | 33 (13.3) | 11 (19.3) | 44 (14.5) |
| | Very good | 93 (37.7) | 24 (42.1) | 117 (38.5) |
| | Good | 73 (29.6) | 13 (22.8) | 86 (28.3) |
| | Fair | 35 (14.2) | 8 (14.0) | 43 (14.1) |
| | Poor | 13 (5.2) | 1 (1.8) | 14 (4.6) |
| Immunodeficiency disorder | | 5 (2.0) | 0 (0.0) | 5 (1.6) |
| Taking systemic immunosuppressant | | 33 (13.4) | 1 (1.8) | 34 (11.2) |
| Primary / booster vaccine regimen | ChAdOx1 / BNT162b2 | 201 (81.4) | 20 (35.1) | 221 (72.7) |
| | ChAdOx1 / mRNA-1273 | 24 (9.7) | 19 (33.3) | 43 (14.1) |
| | BNT162b2 / BNT162b2 | 22 (8.9) | 18 (31.6) | 40 (13.2) |
| Median time from date of booster to date of sampling, weeks (IQR) | | 7.5 (5.7-9.3) | 7.9 (6.0-10.5) | 7.8 (5.8-10.4) |

Abbreviations: IQR, inter-quartile range; s.d., standard deviation; IMD, index of multiple deprivation; Ig, Immunoglobulin.
(1) Ethnicity not reported for n=1 who received BNT162b2 vaccine
(2) Nation of residence not reported for n=1 who received BNT162b2 vaccine and n=2 who received ChAdOx1 vaccine
(3) BMI not reported for n=10 who received BNT162b2 vaccine and n=2 who received ChAdOx1 vaccine
(4) Level of education not reported for n=5 who received BNT162b2 vaccine and n=1 who received ChAdOx1 vaccine
(5) IMD rank not reported for n=2 who received BNT162b2 vaccine and n=5 who received ChAdOx1 vaccine

**Table S4.** Determinants of antibody titres following administration of two doses of ChAdOx1 nCoV-19 vaccine.

| Predictor | N Seropositive (%) | Median IgGAM ratio (IQR) | Minimally Adjusted % difference (95% CI) [1] | P value | Fully Adjusted % difference (95% CI) [2] | P value |
|---|---|---|---|---|---|---|
| Vaccine timing | | | | | | |
| Time since fully vaccinated, wks | | | | | | |
| <2 | 17/5436 (0.3) | 2.31 (1.48, 3.08) | -8.14 (-27.78, 16.85) | 0.49 | -12.25 (-31.35, 12.17) | 0.30 |
| 2-4 | 513/5436 (9.4) | 2.71 (2.01, 3.80) | Referent | | | |
| 5-8 | 2924/5436 (53.8) | 2.45 (1.80, 3.35) | -7.99 (-12.27, -3.51) | 0.001 | -8.39 (-12.64, -3.95) | <0.001 |
| 9-16 | 1890/5436 (34.8) | 2.22 (1.68, 2.99) | -14.78 (-19, -10.34) | <0.001 | -13.67 (-18, -9.1) | <0.001 |
| >16 | 92/5436 (1.7) | 2.39 (1.66, 3.77) | -9.11 (-18.69, 1.6) | 0.09 | 3.04 (-9.36, 17.13) | 0.65 |
| Inter-vaccine interval, weeks | | | | | | |
| <6 | 110/5436 (2.0) | 1.92 (1.38, 2.98) | -14.8 (-22.5, -6.34) | 0.001 | -10.2 (-19.66, 0.38) | 0.06 |
| 6-10 | 1731/5436 (31.8) | 2.34 (1.71, 3.17) | -6.27 (-8.94, -3.52) | <0.001 | -4.36 (-7.25, -1.38) | 0.004 |
| >10 | 3595/5436 (66.1) | 2.44 (1.81, 3.35) | Referent | | | |
| Time of second vaccination | | | | | | |
| Morning | 2291/5254 (43.6) | 2.38 (1.76, 3.27) | Referent | | | |
| Lunchtime | 930/5254 (17.7) | 2.35 (1.75, 3.18) | -1.93 (-5.6, 1.88) | 0.32 | | |
| Afternoon | 1500/5254 (28.5) | 2.43 (1.78, 3.36) | 1.96 (-1.3, 5.34) | 0.24 | | |
| Evening | 533/5254 (10.1) | 2.42 (1.78, 3.36) | 0.21 (-4.42, 5.06) | 0.93 | | |
| Quarter of second vaccination | | | | | | |
| Q1 | 217/5436 (4.0) | 2.08 (1.66, 2.91) | -9.84 (-15.77, -3.49) | 0.003 | -6.31 (-14.47, 2.62) | 0.16 |
| Q2 | 5204/5436 (95.7) | 2.41 (1.78, 3.31) | Referent | | | |
| Q3 | 9/5436 (0.2) | 1.38 (1.36, 3.24) | -13.65 (-37.7, 19.68) | 0.38 | 33.63 (-10.87, 100.34) | 0.16 |
| Q4 | 6/5436 (0.1) | 1.28 (1.22, 1.49) | -44.55 (-62.82, -17.3) | 0.004 | -72.2 (-85.48, -46.78) | <0.001 |
| Socio-demographic factors (baseline) | | | | | | |
| Age | | | | | | |
| <30 | 27/5436 (0.5) | 2.34 (1.60, 3.69) | Referent | | | |
| 30-39.99 | 92/5436 (1.7) | 2.42 (1.79, 3.15) | 2.02 (-17.61, 26.34) | 0.85 | | |
| 40-49.99 | 350/5436 (6.4) | 2.47 (1.88, 3.51) | 9.83 (-9.63, 33.49) | 0.35 | | |
| 50-59.99 | 1519/5436 (27.9) | 2.61 (1.91, 3.59) | 14.13 (-5.58, 37.96) | 0.17 | | |
| 60-69.99 | 2358/5436 (43.4) | 2.37 (1.75, 3.21) | 2.65 (-15.03, 24.01) | 0.79 | | |
| 70-79.99 | 1052/5436 (19.4) | 2.18 (1.64, 2.97) | -4.22 (-20.83, 15.88) | 0.66 | | |
| ≥80 | 38/5436 (0.7) | 1.83 (1.39, 2.44) | -16.61 (-34.79, 6.65) | 0.15 | | |
| Sex | | | | | | |
| Female | 3895/5436 (71.7) | 2.41 (1.79, 3.32) | Referent | | | |
| Male | 1541/5436 (28.3) | 2.34 (1.71, 3.21) | -1.66 (-4.54, 1.32) | 0.27 | | |
| Ethnicity | | | | | | |

| Predictor | N Seropositive (%) | Median IgGAM ratio (IQR) | Minimally Adjusted % difference (95% CI) [1] | P value | Fully Adjusted % difference (95% CI) [2] | P value |
|---|---|---|---|---|---|---|
| White | 5236/5436 (96.3) | 2.38 (1.76, 3.26) | Referent | | | |
| Mixed/Multiple/Other ethnic group | 116/5436 (2.1) | 2.67 (1.97, 3.73) | 17.1 (6.83, 28.36) | 0.001 | 10.19 (-0.01, 21.44) | 0.05 |
| South Asian | 61/5436 (1.1) | 3.15 (2.22, 4.06) | 20.78 (6.46, 37.02) | 0.003 | 21.23 (4.64, 40.45) | 0.01 |
| Black/African/Caribbean/Black British | 23/5436 (0.4) | 2.98 (2.20, 5.12) | 27.16 (3.65, 56.01) | 0.021 | 10.44 (-11.26, 37.43) | 0.37 |
| BMI, kg/m2 | | | | | | |
| <25 | 2702/5427 (49.8) | 2.38 (1.77, 3.20) | Referent | | | |
| 25-30 | 1758/5427 (32.4) | 2.38 (1.76, 3.28) | 2.74 (-0.31, 5.88) | 0.079 | 1.96 (-1.22, 5.24) | 0.23 |
| >30 | 967/5427 (17.8) | 2.49 (1.74, 3.61) | 4.68 (0.90, 8.60) | 0.015 | 3.63 (-0.54, 7.97) | 0.09 |
| Highest educational level attained | | | | | | |
| Primary/Secondary | 587/5435 (10.8) | 2.34 (1.66, 3.30) | 3.62 (-1.16, 8.63) | 0.14 | 0.98 (-3.97, 6.18) | 0.70 |
| Higher/further (A levels) | 805/5435 (14.8) | 2.52 (1.83, 3.52) | 7.08 (2.66, 11.69) | 0.001 | 3.28 (-1.17, 7.93) | 0.15 |
| College | 2409/5435 (44.3) | 2.43 (1.80, 3.32) | 4.76 (1.53, 8.10) | 0.004 | 3.99 (0.64, 7.45) | 0.02 |
| Post-grad | 1634/5435 (30.1) | 2.34 (1.73, 3.17) | Referent | | | |
| Quantiles of IMD rank | | | | | | |
| Q1 (least wealthy) | 1093/5431 (20.1) | 2.43 (1.78, 3.42) | 1.97 (-1.90, 6.00) | 0.35 | | |
| Q2 | 1335/5431 (24.6) | 2.36 (1.73, 3.22) | -1.49 (-5.03, 2.18) | 0.45 | | |
| Q3 | 1452/5431 (26.7) | 2.39 (1.79, 3.28) | -0.11 (-3.62, 3.53) | 0.95 | | |
| Q4 (most wealthy) | 1551/5431 (28.6) | 2.39 (1.77, 3.28) | Referent | | | |
| Lifestyle factors | | | | | | |
| Tobacco smoking | | | | | | |
| No | 5223/5436 (96.1) | 2.39 (1.76, 3.29) | Referent | | | |
| Yes | 213/5436 (3.9) | 2.52 (1.88, 3.42) | 0.77 (-5.92, 7.94) | 0.83 | | |
| Vaping | | | | | | |
| No | 5304/5422 (97.8) | 2.39 (1.76, 3.29) | Referent | | | |
| Yes | 118/5422 (2.2) | 2.65 (1.92, 3.63) | 7.87 (-1.55, 18.20) | 0.10 | | |
| Alcohol, units/wk | | | | | | |
| None | 1403/5436 (25.8) | 2.42 (1.77, 3.26) | Referent | | | |
| 1-7 | 1924/5436 (35.4) | 2.41 (1.79, 3.35) | 1.75 (-1.69, 5.31) | 0.32 | | |
| 8-14 | 1113/5436 (20.5) | 2.33 (1.73, 3.25) | 0.70 (-3.20, 4.75) | 0.73 | | |
| 15-21 | 554/5436 (10.2) | 2.40 (1.76, 3.32) | 2.50 (-2.44, 7.69) | 0.33 | | |
| 22-28 | 249/5436 (4.6) | 2.22 (1.73, 2.96) | -3.95 (-10.25, 2.80) | 0.25 | | |
| >28 | 193/5436 (3.6) | 2.48 (1.82, 3.45) | 3.38 (-4.16, 11.52) | 0.39 | | |
| Light exercise, hrs/wk | | | | | | |
| 0-4 | 1643/5420 (30.3) | 2.47 (1.81, 3.45) | 3.91 (0.52, 7.42) | 0.024 | 1.78 (-1.76, 5.45) | 0.33 |
| 5-9 | 1785/5420 (32.9) | 2.37 (1.73, 3.27) | 0.82 (-2.36, 4.10) | 0.62 | -0.54 (-3.84, 2.88) | 0.76 |
| ≥10 | 1992/5420 (36.8) | 2.36 (1.76, 3.19) | Referent | | | |
| Vigorous exercise, hrs/wk | | | | | | |

| Predictor | N Seropositive (%) | Median IgGAM ratio (IQR) | Minimally Adjusted % difference (95% CI) [1] | P value | Fully Adjusted % difference (95% CI) [2] | P value |
|---|---|---|---|---|---|---|
| 0 | 2032/5420 (37.5) | 2.42 (1.76, 3.39) | 4.87 (1.33, 8.54) | 0.007 | 1.76 (-2.01, 5.67) | 0.37 |
| 1-3 | 2013/5420 (37.1) | 2.39 (1.78, 3.28) | 2.91 (-0.57, 6.51) | 0.10 | 1.08 (-2.47, 4.77) | 0.56 |
| ≥4 | 1375/5420 (25.4) | 2.36 (1.73, 3.15) | Referent | | | |
| Sleep, hrs/night | | | | | | |
| ≤5 | 468/5433 (8.6) | 2.52 (1.88, 3.48) | 4.83 (-0.51, 10.44) | 0.099 | 3.44 (-2.18, 9.39) | 0.24 |
| 6 | 1301/5433 (23.9) | 2.37 (1.78, 3.31) | -0.42 (-4.09, 3.40) | 0.74 | -1.37 (-5.18, 2.58) | 0.49 |
| 7 | 2242/5433 (41.3) | 2.37 (1.73, 3.24) | -2.31 (-5.50, 0.99) | 0.19 | -1.66 (-4.99, 1.79) | 0.34 |
| ≥8 | 1422/5433 (26.2) | 2.41 (1.77, 3.32) | Referent | | | |
| Self-assessed general health | | | | | | |
| Excellent | 1134/5436 (20.9) | 2.38 (1.78, 3.23) | Referent | | | |
| Very good | 2215/5436 (40.7) | 2.37 (1.73, 3.24) | 0.38 (-3.15, 4.04) | 0.84 | -0.18 (-3.82, 3.6) | 0.93 |
| Good | 1350/5436 (24.8) | 2.44 (1.82, 3.37) | 4.25 (0.21, 8.45) | 0.039 | 2.48 (-1.83, 6.98) | 0.26 |
| Fair | 584/5436 (10.7) | 2.43 (1.77, 3.35) | 1.37 (-3.58, 6.56) | 0.6 | 0.97 (-4.54, 6.79) | 0.74 |
| Poor | 153/5436 (2.8) | 2.38 (1.70, 3.65) | 4.91 (-3.59, 14.16) | 0.27 | -2.58 (-11.87, 7.69) | 0.61 |
| Anxiety or depression | | | | | | |
| No | 4096/5433 (75.4) | 2.39 (1.76, 3.27) | Referent | | | |
| Yes | 1337/5433 (24.6) | 2.41 (1.77, 3.35) | 0.08 (-3.00, 3.26) | 0.96 | | |
| Food choice | | | | | | |
| None | 5123/5436 (94.2) | 2.40 (1.77, 3.30) | Referent | | | |
| Vegetarian | 237/5436 (4.4) | 2.34 (1.73, 3.31) | -3.42 (-9.52, 3.10) | 0.30 | | |
| Vegan | 76/5436 (1.4) | 2.33 (1.74, 2.95) | -6.65 (-16.65, 4.55) | 0.23 | | |
| Medical conditions | | | | | | |
| Heart disease [3] | | | | | | |
| No | 5236/5436 (96.3) | 2.40 (1.77, 3.31) | Referent | | | |
| Yes | 200/5436 (3.7) | 2.15 (1.56, 2.75) | -5.04 (-11.61, 2.02) | 0.16 | | |
| Arterial disease [4] | | | | | | |
| No | 5169/5436 (95.1) | 2.40 (1.77, 3.31) | Referent | | | |
| Yes | 267/5436 (4.9) | 2.20 (1.59, 2.93) | -4.11 (-9.93, 2.08) | 0.19 | | |
| Hypertension | | | | | | |
| No | 4198/5436 (77.2) | 2.42 (1.80, 3.32) | Referent | | | |
| Yes | 1238/5436 (22.8) | 2.32 (1.68, 3.20) | -1.39 (-4.55, 1.87) | 0.4 | | |
| Immunodeficiency [5] | | | | | | |
| No | 5412/5436 (99.6) | 2.40 (1.77, 3.30) | Referent | | | |
| Yes | 24/5436 (0.4) | 1.95 (1.61, 2.67) | -14.47 (-30.00, 4.52) | 0.13 | | |
| Major neurological conditions [6] | | | | | | |
| No | 5304/5436 (97.6) | 2.40 (1.77, 3.31) | Referent | | | |
| Yes | 132/5436 (2.4) | 1.98 (1.56, 2.88) | -9.84 (-17.31, -1.69) | 0.02 | -8.84 (-16.78, -0.14) | 0.05 |

| Predictor | N Seropositive (%) | Median IgGAM ratio (IQR) | Minimally Adjusted % difference (95% CI) [1] | P value | Fully Adjusted % difference (95% CI) [2] | P value |
|---|---|---|---|---|---|---|
| Cancer | | | | | | |
| Never | 4899/5436 (90.1) | 2.40 (1.78, 3.31) | Referent | | | |
| Past (cured or in remission) | 497/5436 (9.1) | 2.34 (1.71, 3.22) | -0.62 (-5.13, 4.10) | 0.79 | -0.73 (-5.4, 4.18) | 0.77 |
| Present (active) | 40/5436 (0.7) | 1.90 (1.46, 2.76) | -15.79 (-27.93, -1.61) | 0.03 | -15.51 (-29.02, 0.59) | 0.06 |
| Asthma | | | | | | |
| No | 4556/5436 (83.8) | 2.38 (1.75, 3.29) | Referent | | | |
| Yes | 880/5436 (16.2) | 2.46 (1.83, 3.41) | 1.74 (-1.87, 5.49) | 0.35 | | |
| COPD | | | | | | |
| No | 5335/5436 (98.1) | 2.40 (1.76, 3.29) | Referent | | | |
| Yes | 101/5436 (1.9) | 2.37 (1.77, 3.70) | 3.78 (-5.96, 14.53) | 0.46 | | |
| Diabetes types | | | | | | |
| No diabetes | 5028/5430 (92.6) | 2.40 (1.78, 3.30) | Referent | | | |
| Pre-diabetes | 181/5430 (3.3) | 2.34 (1.71, 3.27) | 0.41 (-6.78, 8.16) | 0.91 | | |
| Type 1 diabetes | 33/5430 (0.6) | 2.29 (1.68, 3.17) | -3.87 (-19.00, 14.08) | 0.65 | | |
| Type 2 diabetes | 188/5430 (3.5) | 2.14 (1.46, 3.14) | -4.14 (-10.9, 3.13) | 0.26 | | |
| Atopy [7] | | | | | | |
| No | 4048/5436 (74.5) | 2.38 (1.76, 3.26) | Referent | | | |
| Yes | 1388/5436 (25.5) | 2.42 (1.80, 3.35) | 0.82 (-2.23, 3.95) | 0.6 | | |
| Pre-vaccination SARS-COV-2 infection/COVID-19 | | | | | | |
| Seronegative | 3491/4150 (84.1) | 2.29 (1.72, 3.02) | Referent | | | |
| Seropositive, asymptomatic | 489/4150 (11.8) | 3.08 (2.16, 4.43) | 40.16 (34.16, 46.42) | <0.001 | 39.83 (33.87, 46.05) | <0.001 |
| Seropositive, symptomatic | 170/4150 (4.1) | 5.02 (3.76, 8.23) | 120.94 (106.67, 136.19) | <0.001 | 119.95 (105.74, 135.14) | <0.001 |
| Nutritional supplements | | | | | | |
| Multivitamin | | | | | | |
| No | 4298/5436 (79.1) | 2.40 (1.76, 3.31) | Referent | | | |
| Yes | 1138/5436 (20.9) | 2.38 (1.78, 3.23) | -1.97 (-5.13, 1.29) | 0.23 | | |
| Vitamin A | | | | | | |
| No | 5407/5436 (99.5) | 2.39 (1.76, 3.30) | Referent | | | |
| Yes | 29/5436 (0.5) | 2.18 (1.73, 3.37) | -8.97 (-24.15, 9.24) | 0.31 | | |
| Vitamin C | | | | | | |
| No | 4887/5436 (89.9) | 2.38 (1.76, 3.28) | Referent | | | |
| Yes | 549/5436 (10.1) | 2.49 (1.81, 3.49) | 5.78 (1.22, 10.55) | 0.01 | 3.28 (-1.45, 8.22) | 0.18 |
| Vitamin D | | | | | | |
| No | 2642/5436 (48.6) | 2.40 (1.78, 3.30) | Referent | | | |
| Yes | 2794/5436 (51.4) | 2.39 (1.75, 3.29) | -0.82 (-3.43, 1.86) | 0.54 | | |

| Predictor | N Seropositive (%) | Median IgGAM ratio (IQR) | Minimally Adjusted % difference (95% CI) [1] | P value | Fully Adjusted % difference (95% CI) [2] | P value |
|---|---|---|---|---|---|---|
| Zinc | | | | | | |
| No | 5189/5436 (95.5) | 2.40 (1.76, 3.30) | Referent | | | |
| Yes | 247/5436 (4.5) | 2.32 (1.78, 3.16) | -3.19 (-9.17, 3.19) | 0.32 | | |
| Selenium | | | | | | |
| No | 5375/5436 (98.9) | 2.40 (1.77, 3.30) | Referent | | | |
| Yes | 61/5436 (1.1) | 2.19 (1.55, 2.99) | -12.98 (-23.29, -1.28) | 0.03 | -16.87 (-27.57, -4.58) | 0.01 |
| Iron | | | | | | |
| No | 5255/5436 (96.7) | 2.40 (1.77, 3.30) | Referent | | | |
| Yes | 181/5436 (3.3) | 2.19 (1.74, 3.18) | -4.79 (-11.60, 2.55) | 0.20 | | |
| Probiotics | | | | | | |
| No | 5078/5436 (93.4) | 2.39 (1.76, 3.30) | Referent | | | |
| Yes | 358/5436 (6.6) | 2.44 (1.83, 3.26) | -0.01 (-5.24, 5.52) | 1.0 | | |
| Omega-3 | | | | | | |
| No | 4765/5436 (87.7) | 2.39 (1.75, 3.29) | Referent | | | |
| Yes | 671/5436 (12.3) | 2.42 (1.85, 3.33) | 3.04 (-1.04, 7.29) | 0.15 | | |
| Cod liver oil | | | | | | |
| No | 4951/5436 (91.1) | 2.40 (1.77, 3.30) | Referent | | | |
| Yes | 485/5436 (8.9) | 2.32 (1.70, 3.17) | -0.86 (-5.40, 3.89) | 0.72 | | |
| Garlic | | | | | | |
| No | 5312/5436 (97.7) | 2.39 (1.76, 3.29) | Referent | | | |
| Yes | 124/5436 (2.3) | 2.60 (1.86, 3.58) | 10.33 (0.92, 20.60) | 0.03 | 15.18 (5.13, 26.2) | 0.002 |
| Medications | | | | | | |
| Beta-2 adrenergic agonists | | | | | | |
| No | 4948/5436 (91.0) | 2.39 (1.76, 3.29) | Referent | | | |
| Yes | 488/5436 (9.0) | 2.49 (1.81, 3.46) | 2.60 (-2.07, 7.49) | 0.28 | | |
| Beta blockers | | | | | | |
| No | 5057/5436 (93.0) | 2.40 (1.78, 3.31) | Referent | | | |
| Yes | 379/5436 (7.0) | 2.29 (1.62, 3.05) | -4.57 (-9.45, 0.58) | 0.08 | -1.95 (-7.33, 3.74) | 0.49 |
| Statins | | | | | | |
| No | 4513/5436 (83.0) | 2.43 (1.80, 3.34) | Referent | | | |
| Yes | 923/5436 (17.0) | 2.18 (1.61, 2.95) | -6.05 (-9.49, -2.48) | 0.001 | -6.57 (-10.24, -2.75) | 0.001 |
| ACE inhibitors | | | | | | |
| No | 4906/5436 (90.3) | 2.41 (1.79, 3.31) | Referent | | | |
| Yes | 530/5436 (9.7) | 2.18 (1.60, 3.08) | -5.62 (-9.81, -1.24) | 0.01 | -5.3 (-9.82, -0.55) | 0.03 |
| Proton pump inhibitors | | | | | | |
| No | 4694/5436 (86.4) | 2.41 (1.78, 3.31) | Referent | | | |
| Yes | 742/5436 (13.6) | 2.26 (1.66, 3.09) | -3.83 (-7.50, -0.02) | 0.05 | -6.33 (-10.25, -2.24) | 0.003 |

| Predictor | N Seropositive (%) | Median IgGAM ratio (IQR) | Minimally Adjusted % difference (95% CI) [1] | P value | Fully Adjusted % difference (95% CI) [2] | P value |
|---|---|---|---|---|---|---|
| H2-receptor antagonists | | | | | | |
| No | 5399/5436 (99.3) | 2.39 (1.76, 3.30) | Referent | | | |
| Yes | 37/5436 (0.7) | 2.38 (1.73, 3.17) | 3.45 (-11.98, 21.60) | 0.68 | | |
| Inhaled corticosteroids | | | | | | |
| No | 5087/5436 (93.6) | 2.40 (1.76, 3.31) | Referent | | | |
| Yes | 349/5436 (6.4) | 2.38 (1.79, 3.15) | -2.38 (-7.53, 3.06) | 0.38 | | |
| Bronchodilators | | | | | | |
| No | 4930/5436 (90.7) | 2.38 (1.76, 3.28) | Referent | | | |
| Yes | 506/5436 (9.3) | 2.51 (1.82, 3.45) | 3.27 (-1.35, 8.11) | 0.17 | | |
| Systemic Immunosuppressants | | | | | | |
| No | 5185/5436 (95.4) | 2.40 (1.76, 3.31) | Referent | | | |
| Yes | 251/5436 (4.6) | 2.36 (1.79, 3.14) | -2.07 (-8.08, 4.33) | 0.52 | | |
| Angiotensin receptor blockers | | | | | | |
| No | 5084/5436 (93.5) | 2.39 (1.77, 3.29) | Referent | | | |
| Yes | 352/5436 (6.5) | 2.39 (1.67, 3.39) | 3.46 (-2.01, 9.25) | 0.22 | | |
| Selective serotonin reuptake inhibitors | | | | | | |
| No | 5104/5436 (93.9) | 2.39 (1.76, 3.28) | Referent | | | |
| Yes | 332/5436 (6.1) | 2.47 (1.77, 3.60) | 4.24 (-1.41, 10.22) | 0.14 | | |
| non-SSRIs antidepressants | | | | | | |
| No | 5218/5436 (96.0) | 2.40 (1.76, 3.29) | Referent | | | |
| Yes | 218/5436 (4.0) | 2.34 (1.74, 3.43) | 1.75 (-4.92, 8.89) | 0.62 | | |
| Calcium channel blockers | | | | | | |
| No | 4870/5436 (89.6) | 2.40 (1.78, 3.31) | Referent | | | |
| Yes | 566/5436 (10.4) | 2.29 (1.65, 3.17) | -1.92 (-6.17, 2.52) | 0.39 | | |
| Thiazides | | | | | | |
| No | 5235/5436 (96.3) | 2.40 (1.77, 3.31) | Referent | | | |
| Yes | 201/5436 (3.7) | 2.22 (1.61, 3.04) | -4.71 (-11.23, 2.29) | 0.18 | | |
| Vitamin K antagonists | | | | | | |
| No | 5399/5436 (99.3) | 2.39 (1.77, 3.30) | Referent | | | |
| Yes | 37/5436 (0.7) | 2.13 (1.72, 3.22) | -2.44 (-17.03, 14.72) | 0.77 | | |
| Sodium-glucose co-transporter-2 inhibitors | | | | | | |
| No | 5414/5436 (99.6) | 2.39 (1.77, 3.30) | Referent | | | |
| Yes | 22/5436 (0.4) | 2.56 (1.43, 4.23) | -1.18 (-19.84, 21.83) | 0.91 | | |
| Anticholinergics | | | | | | |
| No | 5192/5436 (95.5) | 2.39 (1.76, 3.29) | Referent | | | |
| Yes | 244/5436 (4.5) | 2.42 (1.81, 3.32) | 0.81 (-5.46, 7.50) | 0.8 | | |
| Metformin | | | | | | |

| Predictor | N Seropositive (%) | Median IgGAM ratio (IQR) | Minimally Adjusted % difference (95% CI) [1] | P value | Fully Adjusted % difference (95% CI) [2] | P value |
|---|---|---|---|---|---|---|
| No | 5301/5436 (97.5) | 2.39 (1.77, 3.29) | Referent | | | |
| Yes | 135/5436 (2.5) | 2.44 (1.54, 3.69) | 2.58 (-5.84, 11.75) | 0.56 | | |
| Bisphosphonates | | | | | | |
| No | 5331/5436 (98.1) | 2.39 (1.76, 3.30) | Referent | | | |
| Yes | 105/5436 (1.9) | 2.36 (1.79, 3.31) | 2.67 (-6.81, 13.12) | 0.59 | | |
| Anti-platelet drugs | | | | | | |
| No | 5101/5436 (93.8) | 2.40 (1.78, 3.31) | Referent | | | |
| Yes | 335/5436 (6.2) | 2.24 (1.58, 3.05) | -4.14 (-9.38, 1.40) | 0.14 | | |
| Sex hormone therapy | | | | | | |
| No | 4995/5436 (91.9) | 2.39 (1.76, 3.29) | Referent | | | |
| Yes | 441/5436 (8.1) | 2.41 (1.82, 3.33) | -0.31 (-5.12, 4.75) | 0.9 | | |
| Aspirin [8] | | | | | | |
| No | 5168/5436 (95.1) | 2.40 (1.77, 3.31) | Referent | | | |
| Yes | 268/5436 (4.9) | 2.22 (1.59, 3.02) | -3.69 (-9.50, 2.50) | 0.24 | | |
| Paracetamol [8] | | | | | | |
| No | 5211/5436 (95.9) | 2.40 (1.77, 3.30) | Referent | | | |
| Yes | 225/5436 (4.1) | 2.33 (1.73, 3.31) | -0.31 (-6.75, 6.57) | 0.93 | | |
| BCG vaccinated | | | | | | |
| No | 633/5427 (11.7) | 2.38 (1.79, 3.13) | Referent | | | |
| Yes | 4285/5427 (79.0) | 2.41 (1.76, 3.32) | 2.09 (-2.09, 6.44) | 0.91 | | |

Abbreviations: IQR, inter quartile range; CI, confidence interval; BMI, body mass index; IMD, index of multiple deprivation; COPD, chronic obstructive pulmonary disease; SARS-COV-2, severe acute respiratory syndrome coronavirus 2; COVID-19, coronavirus disease 2019; ACE, angiotensin-converting enzyme; H2, histamine 2; SSRIs, selective serotonin reuptake inhibitors; BCG, bacillus calmette-guérin

(1) Adjusted for age and sex only
(2) Adjusted for age, sex, Time since fully vaccinated, inter-dose interval, quarter of second vaccination, ethnicity, BMI, level of education, light exercise, vigorous exercise, sleep, self-assessed general health, major neurological condition, cancer, pre-vaccination SARS-CoV-2 status, use of vitamin C supplements, selenium supplements, garlic supplements, beta blockers, statins, ACE inhibitors, and proton pump inhibitors.
(3) Heart disease defined as coronary artery disease or heart failure
(4) Arterial disease defined as ischaemic heart disease, peripheral vascular disease or cerebrovascular disease
(5) Immunodeficiency defined as HIV, primary immune deficiency or other immunodeficiency
(6) Major neurological conditions defined as stroke, transient ischaemic attack, dementia, Parkinson's disease, multiple sclerosis or motor neuron disease
(7) Atopy defined as atopic eczema/dermatitis and/or hayfever/allergic rhinitis
(8) Chronic use prior to vaccination (i.e. distinct from acute post-vaccination use for treatment of reactogenic symptoms)

**Table S5.** Determinants of antibody titres following administration of two doses of BNT162b2 vaccine.

| Predictor | N Seropositive (%) | Median IgGAM ratio (IQR) | Minimally Adjusted % difference (95% CI) [1] | P value | Fully Adjusted % difference (95% CI) [2] | P value |
|---|---|---|---|---|---|---|
| Vaccine timing | | | | | | |
| Time since fully vaccinated, wks | | | | | | |
| <2 | 14/3291 (0.4) | 10.56 (2.96, 14.2 | 46.59 (9.91, 95.50) | 0.009 | 58.19 (3.5, 141.77) | 0.03 |
| 2-4 | 61/3291 (1.9) | 4.35 (3.32, 5.98) | Referent | | | |
| 5-8 | 827/3291 (25.1) | 4.20 (3.55, 4.96) | -1.85 (-13.72, 11.66) | 0.78 | -10.27 (-22.68, 4.14) | 0.15 |
| 9-16 | 2052/3291 (62.4) | 3.89 (3.14, 4.82) | -13.01 (-23.33, -1.30) | 0.031 | -17.34 (-28.73, -4.14) | 0.01 |
| >16 | 337/3291 (10.2) | 3.26 (2.21, 4.59) | -29.31 (-38.25, -19.07) | <0.001 | -19.76 (-32.85, -4.12) | 0.02 |
| Inter-vaccine interval, weeks | | | | | | |
| <6 | 339/3291 (10.3) | 3.21 (2.32, 4.35) | -23.39 (-27.7, -18.84) | <0.001 | -12.28 (-20.34, -3.41) | 0.01 |
| 6-10 | 1086/3291 (33.0) | 3.85 (3.10, 4.75) | -10.29 (-13.6, -6.86) | <0.001 | -6.32 (-10.67, -1.75) | 0.01 |
| >10 | 1866/3291 (56.7) | 4.11 (3.35, 5.00) | Referent | | | |
| Time of second vaccination | | | | | | |
| Morning | 1253/3158 (39.7) | 4.00 (3.22, 4.87) | Referent | | | |
| Lunchtime | 557/3158 (17.6) | 3.97 (3.21, 4.84) | -2.72 (-7.48, 2.27) | 0.28 | -7.72 (3.59, -55.5) | 0.45 |
| Afternoon | 970/3158 (30.7) | 4.00 (3.10, 4.90) | -1.14 (-5.21, 3.11) | 0.59 | -5.31 (4.19, -21.9) | 0.78 |
| Evening | 378/3158 (12.0) | 3.76 (2.90, 4.80) | -6.75 (-11.99, -1.2) | 0.018 | -9.37 (3.96, -60.5) | 0.40 |
| Quarter of second vaccination | | | | | | |
| Q1 | 1045/3291 (31.8) | 3.66 (2.79, 4.63) | -14.33 (-17.43, -11.11) | <0.001 | -7.12 (-12.84, -1.03) | 0.02 |
| Q2 | 2238/3291 (68.0) | 4.09 (3.33, 4.97) | Referent | | | |
| Q3 | 2/3291 (0.1) | 13.03 (4.47, 21.5 | 126.62 (13.39, 352.89) | 0.021 | - | |
| Q4 | 6/3291 (0.2) | 3.24 (2.88, 4.32) | -17.43 (-44.67, 23.23) | 0.35 | 36.15 (-44.77, 235.66) | 0.50 |
| Socio-demographic factors | | | | | | |
| Age | | | | | | |
| <30 | 53/3291 (1.6) | 4.26 (3.50, 6.09) | Referent | | | |
| 30-39.99 | 94/3291 (2.9) | 4.44 (3.60, 5.28) | -7 (-21.5, 10.18) | 0.4 | -14.01 (-31.27, 7.58) | 0.19 |
| 40-49.99 | 234/3291 (7.1) | 4.05 (3.29, 5.11) | -12.82 (-24.97, 1.3) | 0.073 | -15.54 (-31, 3.38) | 0.10 |
| 50-59.99 | 685/3291 (20.8) | 4.18 (3.40, 5.19) | -8.3 (-20.33, 5.55) | 0.227 | -13.81 (-28.73, 4.23) | 0.13 |
| 60-69.99 | 1196/3291 (36.3) | 3.99 (3.23, 4.87) | -13.62 (-24.79, -0.78) | 0.038 | -19.3 (-33.18, -2.55) | 0.03 |
| 70-79.99 | 894/3291 (27.2) | 3.76 (3.00, 4.59) | -19.33 (-29.85, -7.24) | 0.003 | -23.05 (-36.36, -6.95) | 0.01 |
| ≥80 | 135/3291 (4.1) | 3.08 (2.34, 3.89) | -34.11 (-43.89, -22.63) | <0.001 | -33.63 (-48.51, -14.46) | 0.002 |
| Sex | | | | | | |
| Female | 2345/3291 (71.3) | 4.03 (3.23, 4.95) | Referent | | | |
| Male | 946/3291 (28.7) | 3.82 (2.92, 4.61) | -4.80 (-8.46, -0.99) | 0.01 | -2.53 (-6.95, 2.09) | 0.28 |
| Ethnicity | | | | | | |

| Predictor | N Seropositive (%) | Median IgGAM ratio (IQR) | Minimally Adjusted % difference (95% CI) [1] | P value | Fully Adjusted % difference (95% CI) [2] | P value |
|---|---|---|---|---|---|---|
| White | 3183/3290 (96.7) | 3.95 (3.13, 4.84) | Referent | | | |
| Mixed/Multiple/Other ethnic group | 59/3290 (1.8) | 4.49 (3.66, 5.71) | 9.38 (-4.02, 24.66) | 0.18 | 9.48 (-6.79, 28.6) | 0.27 |
| South Asian | 38/3290 (1.2) | 4.04 (3.42, 5.25) | 6.03 (-9.82, 24.67) | 0.48 | 14.34 (-5.9, 38.94) | 0.18 |
| Black/African/Caribbean/Black British | 10/3290 (0.3) | 5.01 (3.91, 7.15) | 38.93 (1.57, 90.04) | 0.040 | 13.34 (-21.34, 63.3) | 0.50 |
| BMI, kg/m2 | | | | | | |
| <25 | 1575/3289 (47.9) | 3.89 (3.11, 4.85) | Referent | | | |
| 25-30 | 1084/3289 (33.0) | 3.98 (3.18, 4.87) | 3.28 (-0.69, 7.41) | 0.11 | | |
| >30 | 630/3289 (19.2) | 4.10 (3.22, 4.86) | 1.01 (-3.61, 5.84) | 0.67 | | |
| Highest educational level attained | | | | | | |
| Primary/Secondary | 396/3286 (12.1) | 4.03 (3.18, 4.94) | 4.71 (-1.37, 11.17) | 0.13 | | |
| Higher/further (A levels) | 437/3286 (13.3) | 3.90 (3.12, 4.84) | 0.84 (-4.78, 6.78) | 0.78 | | |
| College | 1481/3286 (45.1) | 3.95 (3.16, 4.88) | 1.34 (-2.73, 5.59) | 0.52 | | |
| Post-grad | 972/3286 (29.6) | 3.99 (3.15, 4.84) | Referent | | | |
| Quantiles of IMD rank | | | | | | |
| Q1 (least wealthy) | 778/3289 (23.7) | 3.89 (3.04, 4.87) | -4.67 (-9.22, 0.1) | 0.055 | -2.74 (-8.26, 3.1) | 0.35 |
| Q2 | 772/3289 (23.5) | 4.09 (3.36, 4.93) | 3.32 (-1.58, 8.48) | 0.19 | 2.3 (-3.37, 8.3) | 0.44 |
| Q3 | 857/3289 (26.1) | 3.84 (3.06, 4.68) | -4.43 (-8.85, 0.21) | 0.061 | -4.93 (-10.01, 0.43) | 0.07 |
| Q4 (most wealthy) | 882/3289 (26.8) | 4.02 (3.17, 4.94) | Referent | | | |
| Lifestyle factors | | | | | | |
| Tobacco smoking | | | | | | |
| No | 3154/3291 (95.8) | 3.97 (3.15, 4.86) | Referent | | | |
| Yes | 137/3291 (4.2) | 3.82 (3.25, 4.88) | -3.06 (-11.10, 5.71) | 0.48 | | |
| Vaping | | | | | | |
| No | 3213/3282 (97.9) | 3.96 (3.15, 4.86) | Referent | | | |
| Yes | 69/3282 (2.1) | 4.13 (3.42, 4.94) | -1.84 (-13.01, 10.76) | 0.76 | | |
| Alcohol, units/wk | | | | | | |
| None | 880/3291 (26.7) | 4.05 (3.24, 5.02) | Referent | | | |
| 1-7 | 1203/3291 (36.6) | 3.95 (3.15, 4.85) | -3.67 (-7.81, 0.65) | 0.095 | -1.85 (-6.76, 3.32) | 0.48 |
| 8-14 | 684/3291 (20.8) | 3.97 (3.14, 4.85) | -4.4 (-9.11, 0.55) | 0.081 | -1.63 (-7.29, 4.37) | 0.59 |
| 15-21 | 309/3291 (9.4) | 3.74 (3.07, 4.61) | -7.85 (-13.72, -1.58) | 0.015 | -3.42 (-10.54, 4.26) | 0.37 |
| 22-28 | 124/3291 (3.8) | 4.04 (3.16, 4.69) | -2.72 (-11.58, 7.02) | 0.57 | -3.26 (-12.92, 7.48) | 0.54 |
| >28 | 91/3291 (2.8) | 3.96 (2.88, 4.97) | -5.33 (-15.2, 5.7) | 0.33 | -3.22 (-14.5, 9.54) | 0.60 |
| Light exercise, hrs/wk | | | | | | |
| 0-4 | 1041/3285 (31.7) | 4.00 (3.18, 4.93) | 1.06 (-3.17, 5.49) | 0.63 | | |
| 5-9 | 1082/3285 (32.9) | 3.95 (3.21, 4.85) | -0.96 (-5.05, 3.29) | 0.65 | | |
| ≥10 | 1162/3285 (35.4) | 3.95 (3.10, 4.84) | Referent | | | |
| Vigorous exercise, hrs/wk | | | | | | |

| Predictor | N Seropositive (%) | Median IgGAM ratio (IQR) | Minimally Adjusted % difference (95% CI) [1] | P value | Fully Adjusted % difference (95% CI) [2] | P value |
|---|---|---|---|---|---|---|
| 0 | 1270/3281 (38.7) | 3.98 (3.17, 4.86) | 1.3 (-3.19, 6) | 0.58 | | |
| 1-3 | 1242/3281 (37.9) | 3.94 (3.15, 4.85) | -0.57 (-5, 4.07) | 0.81 | | |
| ≥4 | 769/3281 (23.4) | 3.97 (3.11, 4.88) | Referent | | | |
| Sleep, hrs/night | | | | | | |
| ≤5 | 286/3291 (8.7) | 4.07 (3.20, 4.96) | 3.06 (-3.72, 10.33) | 0.39 | | |
| 6 | 821/3291 (24.9) | 4.02 (3.22, 4.94) | 0.82 (-4, 5.89) | 0.74 | | |
| 7 | 1375/3291 (41.8) | 3.92 (3.12, 4.80) | -2.89 (-7.06, 1.47) | 0.19 | | |
| ≥8 | 809/3291 (24.6) | 3.93 (3.12, 4.88) | Referent | | | |
| Self-assessed general health | | | | | | |
| Excellent | 651/3291 (19.8) | 3.95 (3.17, 4.91) | Referent | | | |
| Very good | 1300/3291 (39.5) | 3.93 (3.13, 4.81) | -1.38 (-5.96, 3.42) | 0.57 | | |
| Good | 916/3291 (27.8) | 4.00 (3.10, 4.84) | 1.19 (-3.82, 6.46) | 0.65 | | |
| Fair | 337/3291 (10.2) | 4.02 (3.32, 4.94) | 3.71 (-2.96, 10.84) | 0.28 | | |
| Poor | 87/3291 (2.6) | 4.07 (2.91, 5.34) | 5.04 (-6.18, 17.6) | 0.39 | | |
| Anxiety or depression | | | | | | |
| No | 2525/3290 (76.7) | 3.94 (3.12, 4.84) | Referent | | | |
| Yes | 765/3290 (23.3) | 4.02 (3.27, 4.93) | 1.96 (-2.16, 6.25) | 0.36 | | |
| Food choice | | | | | | |
| None | 3122/3291 (94.9) | 3.96 (3.15, 4.85) | Referent | | | |
| Vegetarian | 134/3291 (4.1) | 3.93 (3.22, 5.01) | -2.68 (-10.84, 6.23) | 0.54 | -5.01 (-14.4, 5.41) | 0.33 |
| Vegan | 35/3291 (1.1) | 4.50 (3.42, 5.82) | 16.03 (-1.93, 37.28) | 0.08 | 16.23 (-7.2, 45.59) | 0.19 |
| Medical conditions | | | | | | |
| Heart disease [3] | | | | | | |
| No | 3120/3291 (94.8) | 3.97 (3.15, 4.87) | Referent | | | |
| Yes | 171/3291 (5.2) | 3.87 (3.03, 4.76) | 3.81 (-4.11, 12.38) | 0.36 | | |
| Arterial disease [4] | | | | | | |
| No | 3063/3291 (93.1) | 3.97 (3.15, 4.88) | Referent | | | |
| Yes | 228/3291 (6.9) | 3.87 (3.16, 4.71) | 3.87 (-3.09, 11.33) | 0.28 | | |
| Hypertension | | | | | | |
| No | 2459/3291 (74.7) | 4.01 (3.21, 4.93) | Referent | | | |
| Yes | 832/3291 (25.3) | 3.83 (3.00, 4.70) | -4.92 (-8.75, -0.94) | 0.02 | -6.14 (-10.83, -1.21) | 0.02 |
| Immunodeficiency [5] | | | | | | |
| No | 3266/3291 (99.2) | 3.97 (3.15, 4.86) | Referent | | | |
| Yes | 25/3291 (0.8) | 3.69 (2.99, 4.44) | 0.60 (-17.56, 22.76) | 0.95 | | |
| Major neurological conditions [6] | | | | | | |
| No | 3179/3291 (96.6) | 3.97 (3.15, 4.86) | Referent | | | |
| Yes | 112/3291 (3.4) | 3.88 (3.08, 4.82) | 4.39 (-5.11, 14.84) | 0.38 | | |

| Predictor | N Seropositive (%) | Median IgGAM ratio (IQR) | Minimally Adjusted % difference (95% CI) [1] | P value | Fully Adjusted % difference (95% CI) [2] | P value |
|---|---|---|---|---|---|---|
| Cancer | | | | | | |
| Never | 2899/3291 (88.1) | 3.97 (3.15, 4.85) | Referent | | | |
| Past (cured or in remission) | 351/3291 (10.7) | 3.96 (3.12, 4.93) | 1.93 (-3.68, 7.87) | 0.51 | | |
| Present (active) | 41/3291 (1.2) | 3.68 (3.25, 4.49) | -3.75 (-17.69, 12.56) | 0.63 | | |
| Asthma | | | | | | |
| No | 2793/3291 (84.9) | 3.98 (3.16, 4.86) | Referent | | | |
| Yes | 498/3291 (15.1) | 3.88 (3.09, 4.84) | -4.51 (-9.03, 0.24) | 0.06 | -2.52 (-7.89, 3.16) | 0.38 |
| COPD | | | | | | |
| No | 3206/3291 (97.4) | 3.97 (3.16, 4.86) | Referent | | | |
| Yes | 85/3291 (2.6) | 3.75 (2.75, 4.55) | -3.60 (-13.56, 7.50) | 0.51 | | |
| Diabetes types | | | | | | |
| No diabetes | 2989/3283 (91.0) | 3.98 (3.17, 4.90) | Referent | | | |
| Pre-diabetes | 99/3283 (3.0) | 3.75 (2.78, 4.39) | -5.96 (-15.03, 4.06) | 0.23 | | |
| Type 1 diabetes | 32/3283 (1.0) | 3.80 (2.90, 4.27) | -13.25 (-27.25, 3.44) | 0.11 | | |
| Type 2 diabetes | 163/3283 (5.0) | 3.86 (3.00, 4.80) | -0.75 (-8.38, 7.51) | 0.85 | | |
| Atopy [7] | | | | | | |
| No | 2465/3291 (74.9) | 3.99 (3.15, 4.88) | Referent | | | |
| Yes | 826/3291 (25.1) | 3.91 (3.12, 4.82) | -2.23 (-6.06, 1.74) | 0.27 | | |
| Pre-vaccination SARS-COV-2 infection/COVID-19 | | | | | | |
| Seronegative | 1705/2012 (84.7) | 3.91 (3.15, 4.67) | Referent | | | |
| Seropositive, asymptomatic | 215/2012 (10.7) | 4.68 (3.76, 6.77) | 37.46 (28.71, 46.8) | <0.001 | 36.06 (27.33, 45.39) | <0.001 |
| Seropositive, symptomatic | 92/2012 (4.6) | 7.29 (4.63, 13.66 | 82.25 (66.61, 99.35) | <0.001 | 84.75 (68.42, 102.67) | <0.001 |
| Nutritional supplements | | | | | | |
| Multivitamin | | | | | | |
| No | 2603/3291 (79.1) | 3.94 (3.15, 4.84) | Referent | | | |
| Yes | 688/3291 (20.9) | 4.03 (3.15, 4.93) | 0.11 (-4.06, 4.46) | 0.96 | | |
| Vitamin A | | | | | | |
| No | 3275/3291 (99.5) | 3.96 (3.15, 4.86) | Referent | | | |
| Yes | 16/3291 (0.5) | 3.96 (3.05, 4.96) | 8.61 (-15.24, 39.16) | 0.51 | | |
| Vitamin C | | | | | | |
| No | 2959/3291 (89.9) | 3.96 (3.14, 4.88) | Referent | | | |
| Yes | 332/3291 (10.1) | 4.06 (3.25, 4.79) | 1.59 (-4.07, 7.58) | 0.59 | | |
| Vitamin D | | | | | | |
| No | 1609/3291 (48.9) | 3.98 (3.17, 4.86) | Referent | | | |
| Yes | 1682/3291 (51.1) | 3.94 (3.12, 4.86) | -0.45 (-3.83, 3.04) | 0.8 | | |
| Zinc | | | | | | |

| Predictor | N Seropositive (%) | Median IgGAM ratio (IQR) | Minimally Adjusted % difference (95% CI) [1] | P value | Fully Adjusted % difference (95% CI) [2] | P value |
|---|---|---|---|---|---|---|
| No | 3118/3291 (94.7) | 3.96 (3.15, 4.85) | Referent | | | |
| Yes | 173/3291 (5.3) | 3.94 (3.27, 4.94) | 4.68 (-3.11, 13.10) | 0.25 | | |
| Selenium | | | | | | |
| No | 3256/3291 (98.9) | 3.96 (3.15, 4.85) | Referent | | | |
| Yes | 35/3291 (1.1) | 4.17 (3.41, 5.68) | 9.89 (-7.11, 30.00) | 0.27 | | |
| Iron | | | | | | |
| No | 3193/3291 (97.0) | 3.96 (3.15, 4.86) | Referent | | | |
| Yes | 98/3291 (3.0) | 4.00 (3.23, 4.99) | 0.32 (-9.38, 11.07) | 0.95 | | |
| Probiotics | | | | | | |
| No | 3091/3291 (93.9) | 3.96 (3.15, 4.85) | Referent | | | |
| Yes | 200/3291 (6.1) | 4.01 (3.13, 5.16) | 4.62 (-2.67, 12.45) | 0.22 | | |
| Omega-3 | | | | | | |
| No | 2874/3291 (87.3) | 3.96 (3.15, 4.84) | Referent | | | |
| Yes | 417/3291 (12.7) | 4.00 (3.21, 5.00) | 3.56 (-1.68, 9.08) | 0.19 | | |
| Cod liver oil | | | | | | |
| No | 2987/3291 (90.8) | 3.96 (3.14, 4.86) | Referent | | | |
| Yes | 304/3291 (9.2) | 4.03 (3.21, 4.88) | 6.24 (0.05, 12.80) | 0.05 | 4.18 (-2.96, 11.84) | 0.26 |
| Garlic | | | | | | |
| No | 3231/3291 (98.2) | 3.96 (3.15, 4.86) | Referent | | | |
| Yes | 60/3291 (1.8) | 4.24 (3.42, 5.29) | 10.80 (-2.62, 26.07) | 0.12 | | |
| Medications | | | | | | |
| Beta-2 adrenergic agonists | | | | | | |
| No | 2993/3291 (90.9) | 3.98 (3.16, 4.86) | Referent | | | |
| Yes | 298/3291 (9.1) | 3.80 (3.06, 4.77) | -4.31 (-9.90, 1.63) | 0.15 | | |
| Beta blockers | | | | | | |
| No | 3007/3291 (91.4) | 3.97 (3.16, 4.86) | Referent | | | |
| Yes | 284/3291 (8.6) | 3.82 (3.02, 4.85) | 0.47 (-5.59, 6.93) | 0.88 | | |
| Statins | | | | | | |
| No | 2547/3291 (77.4) | 4.03 (3.20, 4.96) | Referent | | | |
| Yes | 744/3291 (22.6) | 3.74 (3.02, 4.61) | -3.33 (-7.52, 1.05) | 0.13 | | |
| ACE inhibitors | | | | | | |
| No | 2896/3291 (88.0) | 3.97 (3.17, 4.88) | Referent | | | |
| Yes | 395/3291 (12.0) | 3.89 (2.99, 4.74) | -0.55 (-5.76, 4.95) | 0.84 | | |
| Proton pump inhibitors | | | | | | |
| No | 2742/3291 (83.3) | 3.98 (3.15, 4.88) | Referent | | | |
| Yes | 549/3291 (16.7) | 3.89 (3.13, 4.77) | 1.71 (-2.92, 6.56) | 0.48 | | |
| H2-receptor antagonists | | | | | | |

| Predictor | N Seropositive (%) | Median IgGAM ratio (IQR) | Minimally Adjusted % difference (95% CI) [1] | P value | Fully Adjusted % difference (95% CI) [2] | P value |
|---|---|---|---|---|---|---|
| No | 3268/3291 (99.3) | 3.96 (3.15, 4.86) | Referent | | | |
| Yes | 23/3291 (0.7) | 4.04 (3.27, 4.84) | -0.59 (-19.18, 22.28) | 0.96 | | |
| Inhaled corticosteroids | | | | | | |
| No | 3069/3291 (93.3) | 3.96 (3.15, 4.86) | Referent | | | |
| Yes | 222/3291 (6.7) | 3.96 (3.30, 4.90) | -0.78 (-7.38, 6.29) | 0.82 | | |
| Bronchodilators | | | | | | |
| No | 2977/3291 (90.5) | 3.98 (3.15, 4.86) | Referent | | | |
| Yes | 314/3291 (9.5) | 3.84 (3.06, 4.88) | -2.42 (-7.99, 3.49) | 0.41 | | |
| Systemic Immunosuppressants | | | | | | |
| No | 3118/3291 (94.7) | 3.98 (3.18, 4.87) | Referent | | | |
| Yes | 173/3291 (5.3) | 3.61 (2.53, 4.75) | -11.62 (-18.18, -4.53) | 0.002 | -8.81 (-17.39, 0.66) | 0.07 |
| Angiotensin receptor blockers | | | | | | |
| No | 3078/3291 (93.5) | 3.98 (3.17, 4.90) | Referent | | | |
| Yes | 213/3291 (6.5) | 3.69 (2.94, 4.43) | -7.46 (-13.79, -0.66) | 0.03 | -0.24 (-8.76, 9.08) | 0.96 |
| Selective serotonin reuptake inhibitors | | | | | | |
| No | 3079/3291 (93.6) | 3.96 (3.14, 4.85) | Referent | | | |
| Yes | 212/3291 (6.4) | 4.04 (3.32, 4.99) | 1.56 (-5.38, 9.01) | 0.67 | | |
| non-SSRIs antidepressants | | | | | | |
| No | 3156/3291 (95.9) | 3.97 (3.15, 4.86) | Referent | | | |
| Yes | 135/3291 (4.1) | 3.77 (3.21, 4.93) | 1.69 (-6.79, 10.94) | 0.71 | | |
| Calcium channel blockers | | | | | | |
| No | 2916/3291 (88.6) | 3.97 (3.16, 4.90) | Referent | | | |
| Yes | 375/3291 (11.4) | 3.89 (3.04, 4.72) | 0.24 (-5.15, 5.94) | 0.93 | | |
| Thiazides | | | | | | |
| No | 3171/3291 (96.4) | 3.97 (3.15, 4.86) | Referent | | | |
| Yes | 120/3291 (3.6) | 3.80 (3.11, 4.90) | -2.09 (-10.73, 7.39) | 0.65 | | |
| Vitamin K antagonists | | | | | | |
| No | 3258/3291 (99.0) | 3.97 (3.15, 4.87) | Referent | | | |
| Yes | 33/3291 (1.0) | 3.31 (2.78, 3.98) | -15.22 (-28.71, 0.82) | 0.06 | -17.15 (-35.51, 6.43) | 0.14 |
| Sodium-glucose co-transporter-2 inhibitors | | | | | | |
| No | 3271/3291 (99.4) | 3.96 (3.15, 4.86) | Referent | | | |
| Yes | 20/3291 (0.6) | 4.38 (3.93, 5.10) | 13.83 (-8.87, 42.18) | 0.25 | | |
| Anticholinergics | | | | | | |
| No | 3110/3291 (94.5) | 3.97 (3.15, 4.86) | Referent | | | |
| Yes | 181/3291 (5.5) | 3.81 (3.11, 4.80) | -2.09 (-9.23, 5.61) | 0.58 | | |
| Metformin | | | | | | |
| No | 3173/3291 (96.4) | 3.96 (3.15, 4.86) | Referent | | | |

| Predictor | N Seropositive (%) | Median IgGAM ratio (IQR) | Minimally Adjusted % difference (95% CI) [1] | P value | Fully Adjusted % difference (95% CI) [2] | P value |
|---|---|---|---|---|---|---|
| Yes | 118/3291 (3.6) | 4.04 (3.12, 4.86) | 3.50 (-5.71, 13.61) | 0.47 | | |
| Bisphosphonates | | | | | | |
| No | 3219/3291 (97.8) | 3.97 (3.15, 4.86) | Referent | | | |
| Yes | 72/3291 (2.2) | 3.60 (2.48, 4.90) | -6.75 (-17.15, 4.96) | 0.25 | | |
| Anti-platelet drugs | | | | | | |
| No | 3024/3291 (91.9) | 3.98 (3.17, 4.88) | Referent | | | |
| Yes | 267/3291 (8.1) | 3.82 (3.02, 4.75) | 2.46 (-3.99, 9.34) | 0.46 | | |
| Sex hormone therapy | | | | | | |
| No | 3034/3291 (92.2) | 3.96 (3.13, 4.85) | Referent | | | |
| Yes | 257/3291 (7.8) | 4.06 (3.29, 5.22) | 0.99 (-5.39, 7.81) | 0.77 | | |
| Aspirin [8] | | | | | | |
| No | 3084/3291 (93.7) | 3.97 (3.17, 4.87) | Referent | | | |
| Yes | 207/3291 (6.3) | 3.89 (2.89, 4.77) | 1.48 (-5.62, 9.11) | 0.69 | | |
| Paracetamol [8] | | | | | | |
| No | 3121/3291 (94.8) | 3.97 (3.15, 4.86) | Referent | | | |
| Yes | 170/3291 (5.2) | 3.83 (3.05, 4.83) | 2.04 (-5.62, 10.32) | 0.61 | | |
| BCG vaccinated | | | | | | |
| No | 399/3283 (12.2) | 3.89 (3.00, 4.90) | Referent | | | |
| Yes | 2601/3283 (79.2) | 3.99 (3.20, 4.89) | 3.48 (-1.89, 9.15) | 0.8 | | |

Abbreviations: IQR, inter quartile range; CI, confidence interval; BMI, body mass index; IMD, index of multiple deprivation; COPD, chronic obstructive pulmonary disease; SARS-COV-2, severe acute respiratory syndrome coronavirus 2; COVID-19, coronavirus disease 2019; ACE, angiotensin-converting enzyme; H2, histamine 2; SSRIs, selective serotonin reuptake inhibitors; BCG, bacillus calmette-guérin

(1) Adjusted for age and sex only
(2) Adjusted for age, sex, time since fully vaccinated, inter-dose interval, time of second vaccination, quarter of second vaccination, ethnicity, quantiles of IMD, alcohol consumption, food choice, hypertension, asthma, pre-vaccination SARS-CoV-2 status, cod liver oil, systemic immunosuppressants, selective serotonin reuptake inhibitors, angiotensin receptor blockers, and vitamin K antagonists
3) Heart disease defined as coronary artery disease or heart failure
(4) Arterial disease defined as ischaemic heart disease, peripheral vascular disease or cerebrovascular disease
(5) Immunodeficiency defined as HIV, primary immune deficiency or other immunodeficiency
(6) Major neurological conditions defined as stroke, transient ischaemic attack, dementia, Parkinson’s disease, multiple sclerosis or motor neuron disease
(7) Atopy defined as atopic eczema/dermatitis and/or hayfever/allergic rhinitis
(8) Chronic use prior to vaccination (i.e. distinct from acute post-vaccination use for treatment of reactogenic symptoms)

### Table S6. Influence of post-vaccination use of paracetamol/NSAIDs on antibody titres

| Predictor | | Median IgGAM ratio (IQR) | Adjusted for main model predictors,[1] % difference (95% CI) | P value | Additionally adjusted for presence vs. absence of post-vaccination symptoms,[2] % difference (95% CI) | P value |
|---|---|---|---|---|---|---|
| **Use of post-vaccination paracetamol / NSAIDs** | None | 2.91 (1.93, 4.09) | Referent | | Referent | |
| | After 1st or 2nd doses | 2.75 (1.85, 3.93) | 3.38 (0.25, 6.60) | 0.034 | 0.67 (-2.27, 3.69) | 0.66 |
| | After both doses | 2.95 (1.97, 4.28) | 5.86 (2.22, 9.62) | 0.001 | 2.98 (-0.40, 6.49) | 0.09 |

Abbreviations: IQR, inter-quartile range; CI, confidence interval ; NSAID, non-steroidal anti-inflammatory drug

(1) Main model predictors: vaccine type, time from 2nd vaccine dose to sampling, inter-dose interval, quarter of 2nd vaccine dose, age, ethnicity, BMI, hypertension and pre-vaccination COVID-19 status
(2) Post-vaccination symptoms include any of the following: tenderness/soreness at the injection site, headache, fever or muscle aches

**Table S7. Vaccine regimen and clinical characteristics of participants with undetectable anti-S IgGAM after 3 vaccine doses (n=8)**

| Primary / booster vaccine regimen | Underlying condition | Immunosuppressant medication |
|---|---|---|
| ChAdOx1 / BNT162b2 | Giant cell arteritis | Tocilizumab |
| BNT162b2 / BNT162b2 | Solid organ transplant | Tacrolimus, Mycophenolate mofetil |
| BNT162b2 / BNT162b2 | Solid organ transplant | Tacrolimus, Mycophenolate mofetil |
| BNT162b2 / BNT162b2 | Common variable immunodeficiency | - |
| ChAdOx1 / BNT162b2 | Primary biliary cirrhosis | Mycophenolate mofetil |
| BNT162b2 / BNT162b2 | Rheumatoid arthritis | Methotrexate, Rituximab |
| ChAdOx1 / BNT162b2 | Eosinophilic granulomatosis with polyangitis | Rituximab |
| ChAdOx1 / BNT162b2 | Cryoglobulinaemia | Rituximab |

**Figure S1.** Study profile

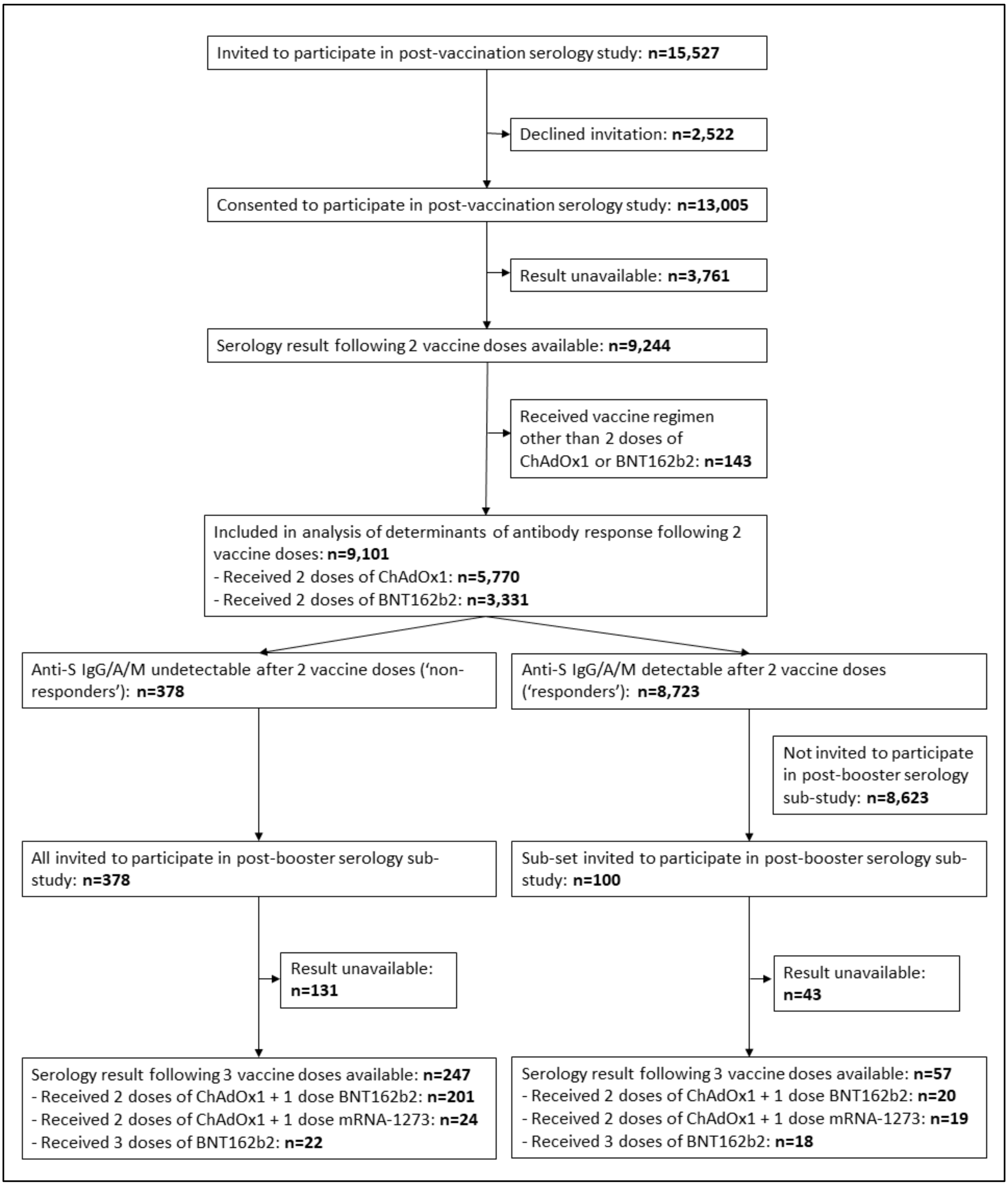

**Figure S2.** Post-vaccination SARS-CoV-2 antibody titres by vaccine type vs. diagnosis of hypertension (A), prescribed immunosuppressant medications (B) or consumption of selenium supplements (C).

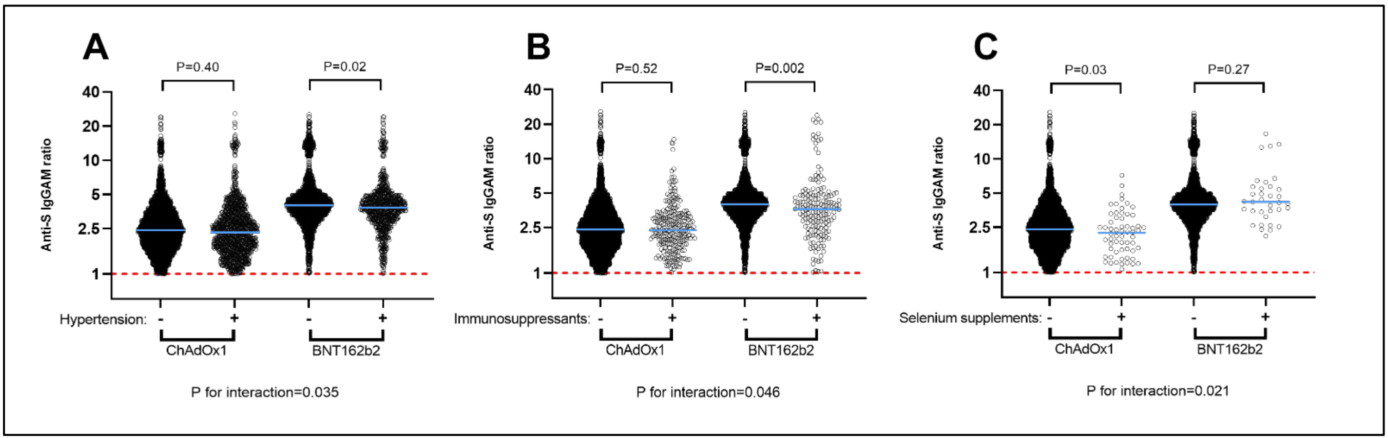
